# Cardiovascular-kidney-metabolic disease burden in children and adults following heart transplantation

**DOI:** 10.1101/2025.02.21.25322699

**Authors:** Shi Huang, Eric Farber-Eger, Jaclyn Tamaroff, Nelson Chow, Hasan K. Siddiqi, D. Marshall Brinkley, Jonathan N. Menachem, Aniket S. Rali, JoAnn Lindenfeld, Henry Ooi, Lynn Punnoose, Stacy Tsai, Dawn Pedrotty, Sandip Zalawadiya, Suzanne Sacks, Mark Wigger, Aaron M. Williams, Swaroop Bommareddi, Brian Lima, Duc Nguyen, Ashish S. Shah, John M. Trahanas, David W. Bearl, Quinn S. Wells, Kelly H. Schlendorf, Kaushik Amancherla

## Abstract

**Background:** Heart transplantation (HT) is the definitive therapy for end-stage heart failure. However, with improving post-HT survival in the modern era, recipients are increasingly cumulatively exposed to unique risk factors for cardiovascular-kidney-metabolic (CKM) dysfunction. An expanded understanding of the incidence and prevalence of CKM dysfunction post-HT may inform screening and therapeutic strategies to mitigate adverse events.

**Objectives:** The aim of this study was to characterize the incidence and prevalence of CKM risk factors in adult and pediatric HT recipients at a high-volume heart transplant center.

**Methods:** We conducted a single-center retrospective observational study in adults and children who underwent HT between 1/1/2015 and 6/30/2024. Longitudinal clinical and laboratory data were extracted from the electronic health record. Incidence rates (IRs) of type 2 diabetes mellitus (DM2), overweight or obesity, hypertension, chronic kidney disease (CKD), and dyslipidemia were calculated. We constructed longitudinal predictive models of CKM dysfunction and evaluated the impact of SGLT2 inhibitors (SGLT2i) and GLP1 receptor agonists (GLP1ra) on CKD and body mass index (BMI), respectively.

**Results:** During the study period, 860 adults and 84 children underwent HT. Among adults, the IR (reported as cases per 100 person-years) of DM2, overweight or obesity, dyslipidemia, and CKD were 28.6, 77.3, 139.1, and 69.7 respectively. Among children, the IR of DM2, overweight or obesity, dyslipidemia, and CKD were 2.8, 26.9, 5.5, and 3.6, respectively. Within 12 months post-HT, 99% of adults developed stage 1 or 2 hypertension and 22.1% of all adults developed HbA1c ≥6.5%, regardless of pre-existing diagnosis of DM2. Similarly, 37.5% of adults developed moderate-severe hypertriglyceridemia and 31.1% manifested worsened LDL-C control. Among adults with an eGFR ≥45 mL/min/1.73 m^2^ pre-HT, 86.2% demonstrated worsening renal function (eGFR <45 mL/min/1.73 m^2^) within 12-months post-HT. In adults initiated on SGLT2i post-HT (N = 242), there was a non-linear improvement in eGFR during the ensuing 12 months while for individuals initiated on GLP1ra (N = 168), there was a predominantly linear reduction in BMI.

**Conclusions:** A significant proportion of HT recipients experience new-onset or worsening CKM dysfunction following HT. Our results indicate that SGLT2i and GLP1ra may mitigate the burden of CKM disease.

## INTRODUCTION

Survival following heart transplantation (HT) has improved steadily over the past few decades^1^. While risk factors and clinical outcomes including acute rejection, cardiac allograft vasculopathy (CAV), and graft survival have been prioritized for study, less studied or understood is the burden of cardiovascular-kidney-metabolic (CKM) disease as HT recipients live increasingly longer. Cardiometabolic disease encompasses type 2 diabetes (DM2), obesity, hypertension, dyslipidemia, and the metabolic syndrome, and has been increasing in prevalence in the United States, leading to increased risk of stroke, cardiovascular disease, and death^2^. Indeed, in recognition of the significant contribution of poor CKM health to morbidity and mortality, the American Heart Association (AHA) has called for improved understanding of major gaps in knowledge of CKM dysfunction^3,4^.

Even prior to transplantation, individuals with end-stage organ disease exhibit a high prevalence of DM2, ranging from 15% to 30%^5^, and obesity^6^. Following HT, recipients are cumulatively exposed to unique risk factors, such as immunosuppressive therapies that directly modulate metabolic pathways and may accelerate metabolic and kidney disease^7–9^. Estimates suggest that within 1-year post-transplant, up to 71% of recipients develop hypertension and 60% develop dyslipidemia^10^. As allograft outcomes are thought to be dependent in part on metabolic risk^11^, the International Society of Heart and Lung Transplantation (ISHLT) recommends frequent screening for both dyslipidemia and DM2^12^. However, granular data on incidence and prevalence of CKM outcomes following HT in adults and children is limited. Furthermore, it remains unclear how novel therapeutic agents, such as SLGT2 inhibitors (SGLT2i) and GLP-1 receptor agonists (GLP1ra), may mitigate CKM dysfunction in this population.

Here, we leverage the extensive clinical data available at our high-volume transplant center to characterize the burden of CKM dysfunction in adult and pediatric HT recipients. Furthermore, in a subset of individuals, we evaluate the impact of SGLT2i on kidney function and GLP1ra on body mass index (BMI). These data inform opportunities to mitigate CKM morbidity following HT.

## METHODS

### Study population

We conducted a retrospective review of all individuals aged ≥ 2 years who underwent HT at Vanderbilt University Medical Center (VUMC) between January 1, 2015 and June 30, 2024. Individuals transplanted elsewhere who transferred their care to VUMC post-HT were included. Individuals referred by the Veterans Affairs system and who underwent HT at VUMC were excluded, as these individuals are not managed at VUMC beyond their index admission. As is the case for all adult HT recipients at our center, maintenance immunosuppression during the first year post-HT consisted of calcineurin inhibitors, antiproliferative agents, and steroids which were progressively weaned off during the first 4-6 months post-HT. Sensitized individuals with panel reactive antibodies > 25% were treated with basiliximab or thymoglobulin induction. Pediatric HT recipients at VUMC undergo thymoglobulin induction followed by a 5-day steroid taper. Subsequently, maintenance therapy in children constitutes a calcineurin inhibitor and antiproliferative agent for 3 months, followed by replacement of the antiproliferative with an mTOR inhibitor (mTORi). This study was approved by the VUMC Institutional Review Board (IRB #200551).

### Data collection and outcomes

Donor data were abstracted from the United Network for Organ Sharing (UNOS) database, while recipient data were extracted from VUMC’s electronic health record (EHR). Data collected included longitudinal laboratory values for BMI, hemoglobin A1c (HbA1c), triglycerides, low-density lipoprotein (LDL-C), cholesterol, high-density lipoprotein (HDL), systolic blood pressure (SBP), diastolic blood pressure (DBP), and estimated glomerular filtration rate (eGFR). In addition, diagnoses (based on ICD-9 or ICD-10 codes) and their respective dates were abstracted as follows: 1) chronic kidney disease (CKD): ICD9 585, 585.*, and ICD10 N18, N18.*; 2) dyslipidemia: ICD9 272.*, and ICD10 E78.* and E88.*; and 3) hypertension: ICD9 codes 401, 401.*, 402, 402.*, 403, 403.*, 404, 404.*, 405, 405.*. ICD10 I10, I10.*, I11, I11.*, I12, I12.*, I13, I13.*, I16, I16.*. Hypertension also was extracted from the EHR when an individual’s problem list included “hypertension” excluding cases where the language also included “portal/pulmonary/rv/intracranial hypertension”. DM2 was defined according to the eMERGE/PheKB definition (https://www.phekb.org/phenotype/type-2-diabetes-mellitus). Longitudinal laboratory data was extracted from the time of HT until 36 months post-HT, and laboratory data was averaged in 3 month increments. We allowed for data collected within ± 30 days of each pre-designated time point post-HT to be included in analyses specific to that time point.

Among adults, BMI was categorized as underweight or healthy weight (BMI < 25 kg/m^2^), overweight (BMI 25 to < 30 kg/m^2^), class I obesity (BMI 30 to < 35 kg/m^2^), class II obesity (BMI 35 to < 40 kg/m^2^), and class III obesity (BMI ≥ 40 kg/m^2^) as defined by the Centers for Disease Control. For children, height and weight were converted to z-scores, followed by calculation of BMI z-scores that were age– and gender-adjusted (https://github.com/CDC-DNPAO/CDCAnthro)^13–15^. BMIs were then categorized as normal weight (BMI z-score <1.04), overweight (BMI z-score of 1.04 to <1.645), and obese (BMI z-score ≥1.645)^14–16^.

In accordance with the American College of Cardiology and AHA guidelines^17^, blood pressure in adults was categorized as normal (SBP < 120 mmHg and DBP < 80 mmHg), elevated (SBP 120-129 mmHg and DBP < 80 mmHg), stage 1 hypertension (SBP 130-139 mmHg or DBP 80-89 mmHg), and stage 2 hypertension (SBP ≥ 140 mmHg or DBP ≥ 90 mmHg). For children, we used the American Academy of Pediatrics guidelines^18^ and assumed 50^th^ percentile for height for simplicity. A normal BP for children aged 2-12 years was defined as < 90^th^ percentile for SBP and DBP. Elevated BP was defined as ≥ 90^th^ percentile to < 95^th^ percentile or 120/80 mmHg to < 95^th^ percentile (whichever was lower). Stage 1 hypertension was defined as ≥ 95^th^ percentile to < 95^th^ percentile + 12 mmHg or 130/80 to 139/89 mmHg (whichever was lower). Finally, stage 2 hypertension was defined as ≥ 95^th^ percentile + 12 mmHg or ≥ 140/90 mmHg, whichever was lower. For children ≥ 13 years of age, normal BP was defined as < 120/80 mmHg, elevated BP as 120/<80 to 129/<80 mmHg, stage 1 hypertension as 130/80 to 139/89 mmHg, and stage 2 hypertension as ≥ 140/90 mmHg. If individuals had multiple BP checks within one day, the average SBP and DBP were used.

Outcomes included incident overweight or obesity, DM2, dyslipidemia, and hypertension following HT, as well as prevalence of these conditions post-HT. In addition, trends in eGFR, HbA1c, SBP, DBP, and lipid profiles across the first 36 months post-HT were examined. For eGFR analyses, we excluded individuals who underwent simultaneous multiorgan transplant as well as those with a history of dialysis pre-HT to minimize any confounding on post-HT eGFR trajectory. For individuals who needed renal replacement therapy (RRT) after HT, we assigned their eGFR value to 0 after their first RRT episode. In a subset of individuals who were prescribed SGLT2i or GLP1ra, we evaluated the impact of these therapies on eGFR and BMI trajectory, respectively, in the 12 months following prescription.

### Statistical analysis

Continuous variables are described as mean ± standard deviation (SD) or median (interquartile range [IQR]). Categorical variables are described as N (%). Kaplan-Meier curves were plotted for incident DM2, worsening HbA1c to ≥ 6.5% (irrespective of previous diagnosis of DM2), dyslipidemia, worsening triglycerides and LDL-C, worsening eGFR, and ≥ stage 1 hypertension. Incident rates (IRs) of disease per 100 person-years were calculated for each variable as follows: [Number of new cases / (Total population at risk x Follow-up time)] x 100. Cumulative incidence of disease burden, estimated by the Kaplan-Meier method, following transplant is reported in percentages (%).

To examine the trajectories of laboratory values over time, trend plots were presented. In addition, to mitigate bias from unequal sample sizes at different time points, a generalized least squares (GLS) model with a first order autoregressive correlation (AR1) was used to construct models of trajectories over 36 months post-HT for the following: HbA1c, triglycerides, LDL-C, cholesterol, HDL, BMI, eGFR, SBP, and DBP. The GLS models include months since HT (0-36) with nonlinear terms with three knots. Similarly, GLS models with AR1 correlation were developed to characterize the trajectories of eGFR and BMI following initiation of SGLT2i and GLP1ra, respectively. Details, including sample sizes at each time point for all CKM variables assessed are outlined explicitly in the **Supplemental Tables**.

We performed sensitivity analyses by repeating all the above analyses, but excluding individuals who died or were lost to follow-up within 30 days of HT. We also performed sensitivity analyses excluding individuals who underwent simultaneous multiorgan transplants. All analyses were performed in R version 4.4.0 (R Core Team).

## RESULTS

### Individual characteristics

During the study period, 860 adults (mean age 52.9 ± 12.8 years at HT) and 84 children (mean age 10.0 ± 4.5 years at HT) underwent HT. Female individuals accounted for 30.1% and 44% of the adult and pediatric cohorts, respectively. Recipient and donor demographics are detailed in **Table 1**.

**Table 1.** Characteristics of adult and pediatric heart transplant recipients.

| **Characteristics (Adults) N = 860<sup>1</sup>** |  | **Characteristics (Children) N = 84<sup>1</sup>** |  |
| --- | --- | --- | --- |
| Age at transplant (years) | 52.9 ± 12.8 | Age at transplant (years) | 10.0 ± 4.5 |
| Recipient gender |  | Recipient gender |  |
| Female | 259 (30.1%) | Female | 37 (44%) |
| Male | 601 (69.9%) | Male | 47 (56%) |
| Ethnicity |  | Ethnicity |  |
| White | 589 (68.5%) | White | 60 (71.4%) |
| Black | 169 (19.7%) | Black | 12 (14.3%) |
| Other | 102 (11.8%) | Other | 12 (14.3%) |
| Deceased | 171 (19.9%) | Deceased | 6 (7.1%) |
| Donor age (years)<sup>2</sup> | 31.0 ± 9.8 | Donor age (years)<sup>3</sup> | 13.7 ± 8.3 |
| Donor gender<sup>2</sup> |  | Donor gender<sup>3</sup> |  |
| Female | 253 (30.6%) | Female | 22 (30.6%) |
| Male | 573 (69.4%) | Male | 50 (69.4%) |

<sup>1</sup>Mean ± SD; n (%)
<sup>2</sup>N = 826
<sup>3</sup>N = 72

### Trends in type 2 diabetes mellitus following heart transplantation in adults

Among 700 adults whose HbA1c was captured at time of HT, mean HbA1c was 5.87% ± 0.86% and steadily increased to a peak of 7.46% ± 2.04% by 30-months post-HT (**Table 2**). Among 434 adults without DM2 prior to HT, based on the KM plot, the cumulative incidence of developing diabetes was 38.8% and 43.8% within 6-months and 1-year post-HT, respectively (**Figure 1A**). The IR of DM2 was 28.6 per 100 person-years (95% CI 24.8-32.4 per 100 person-years). In 507 adults with HbA1c <6.5% prior to HT irrespective of prior DM2 diagnosis, the cumulative incidence of developing worsening HbA1c to ≥6.5% at 6-months post-HT and 1-year post-HT were 12.6% and 22.1%, respectively (**Figure 1B**). The IR of worsening HbA1c following HT was 17.2 per 100 person-years (95% CI 14.6-19.8 per 100 person-years). A GLS model of HbA1c trajectory post-HT suggested time after HT was a significant predictor of HbA1c values (P_chunk_ < 0.001; **Figure 1C**). We found both significant linear (β = 0.049, P-value <0.001) and nonlinear (β = –0.047, P-value = 0.043) time effect to HbA1c levels following HT.

**Figure 1.**
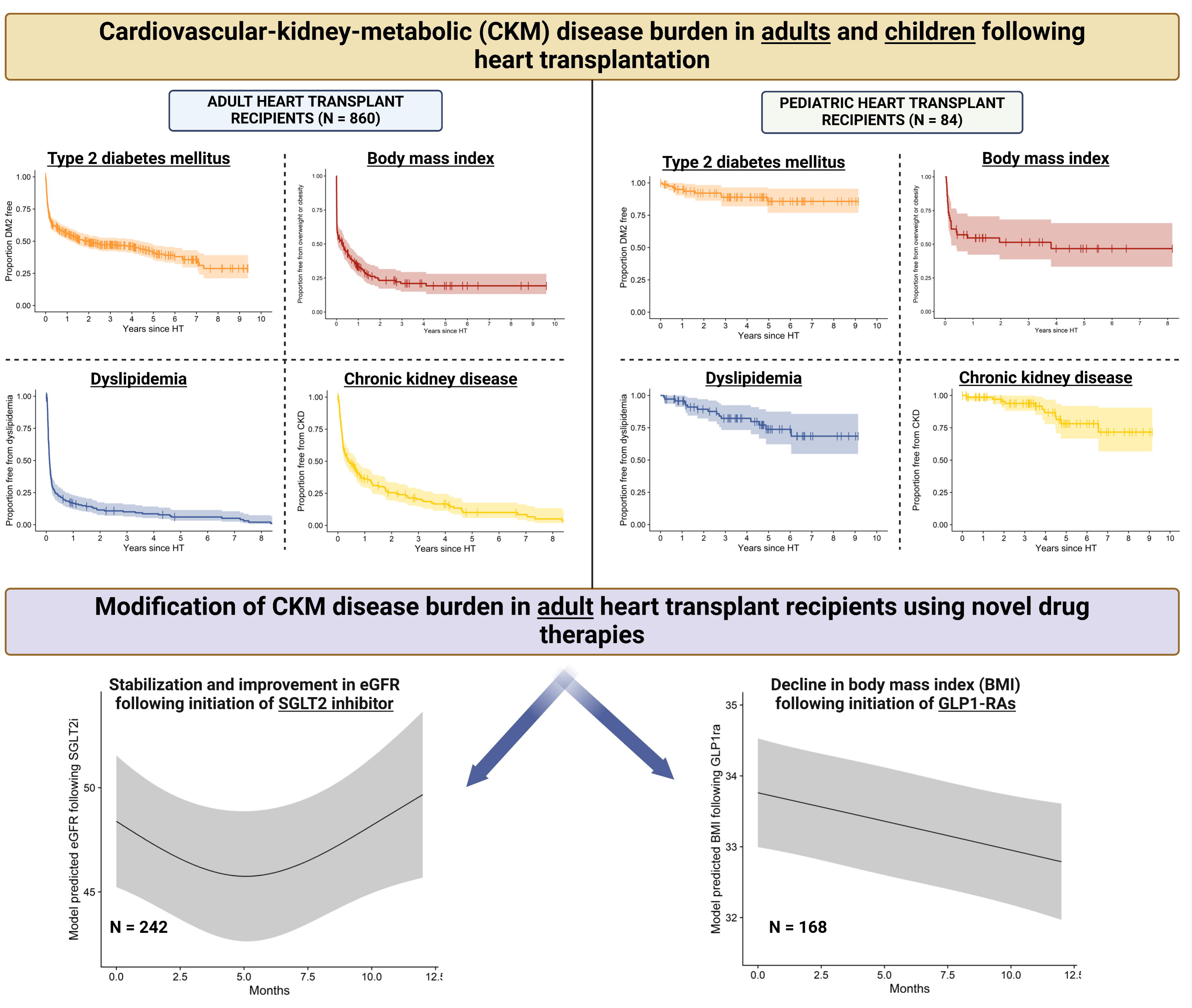
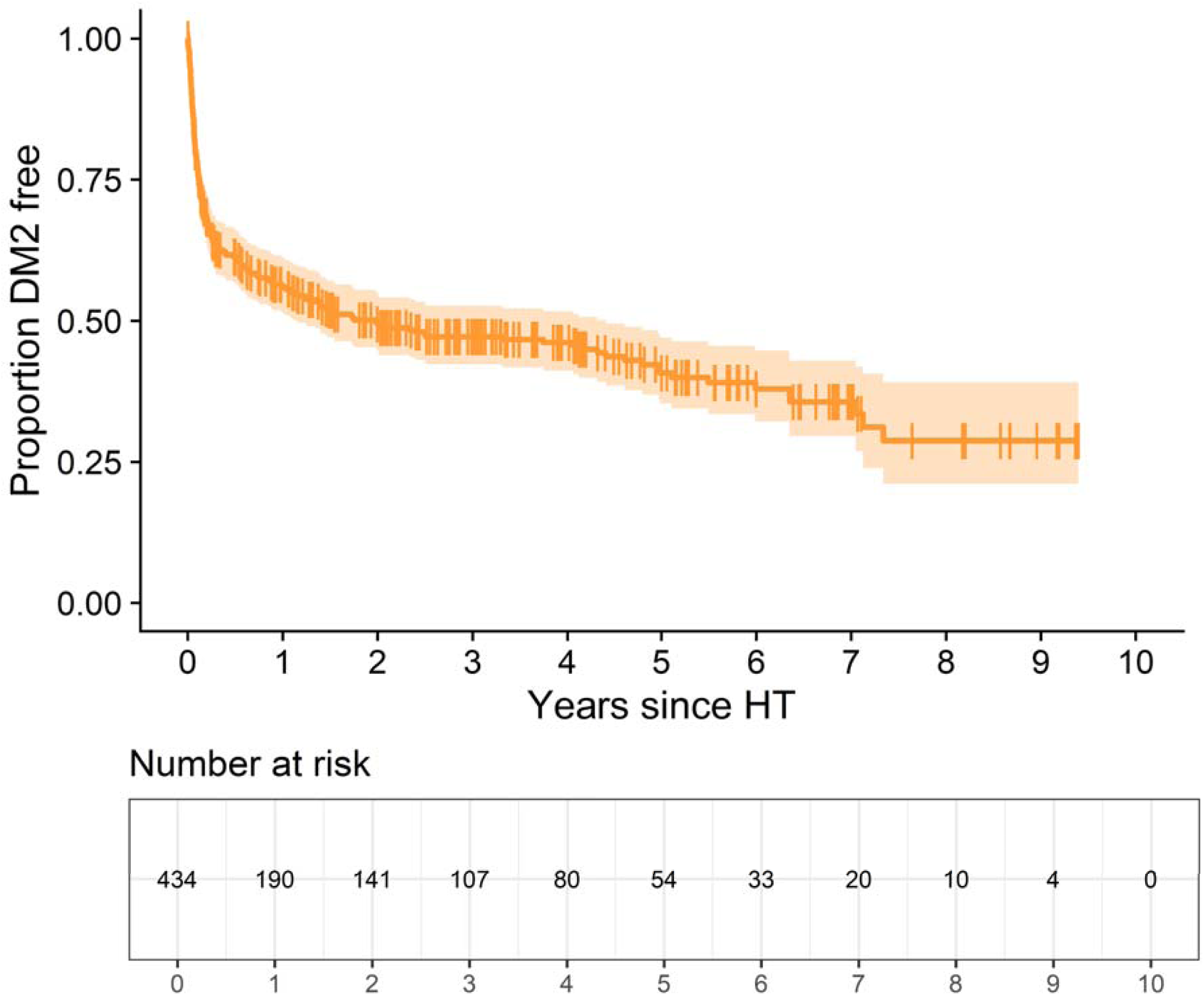
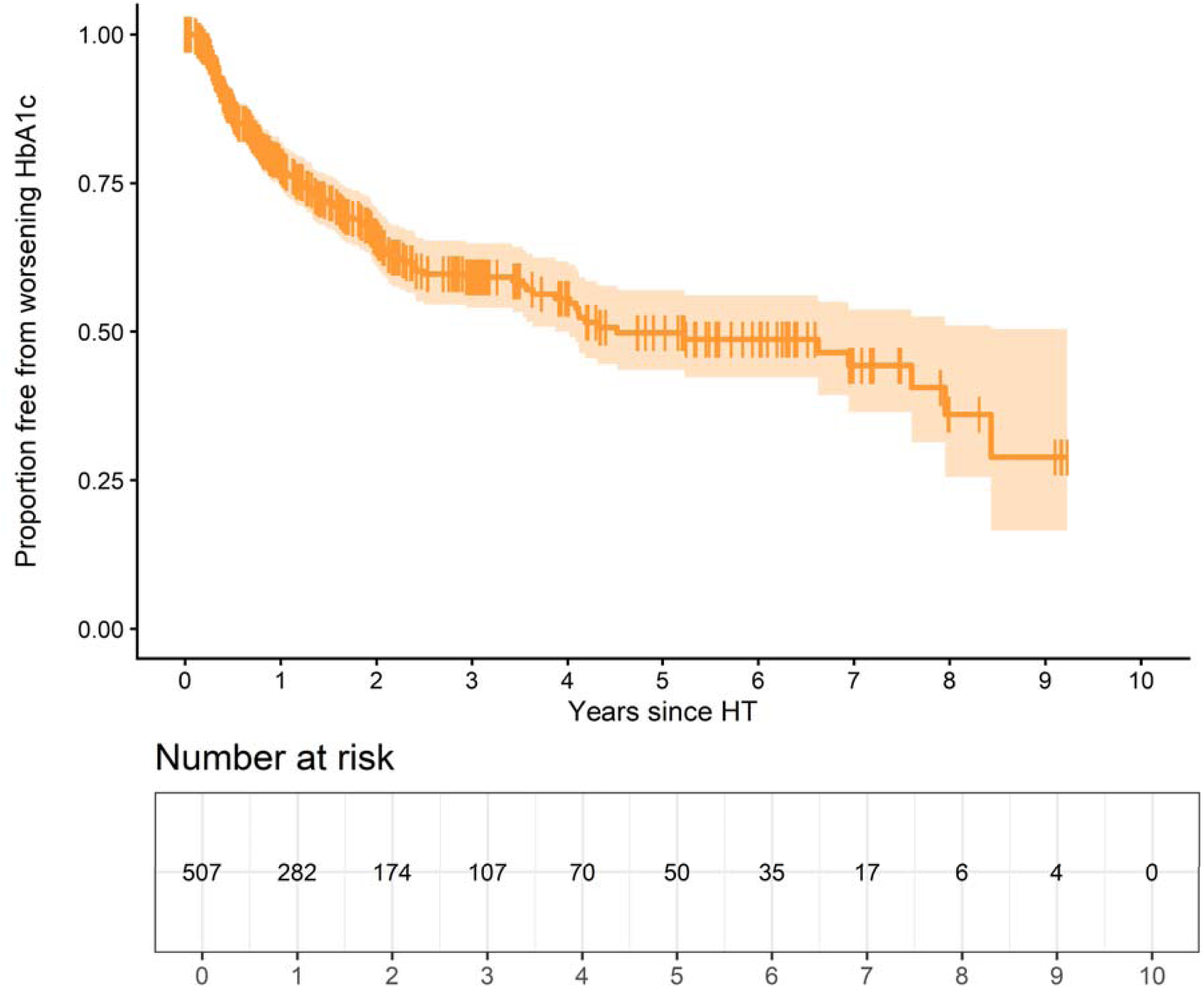
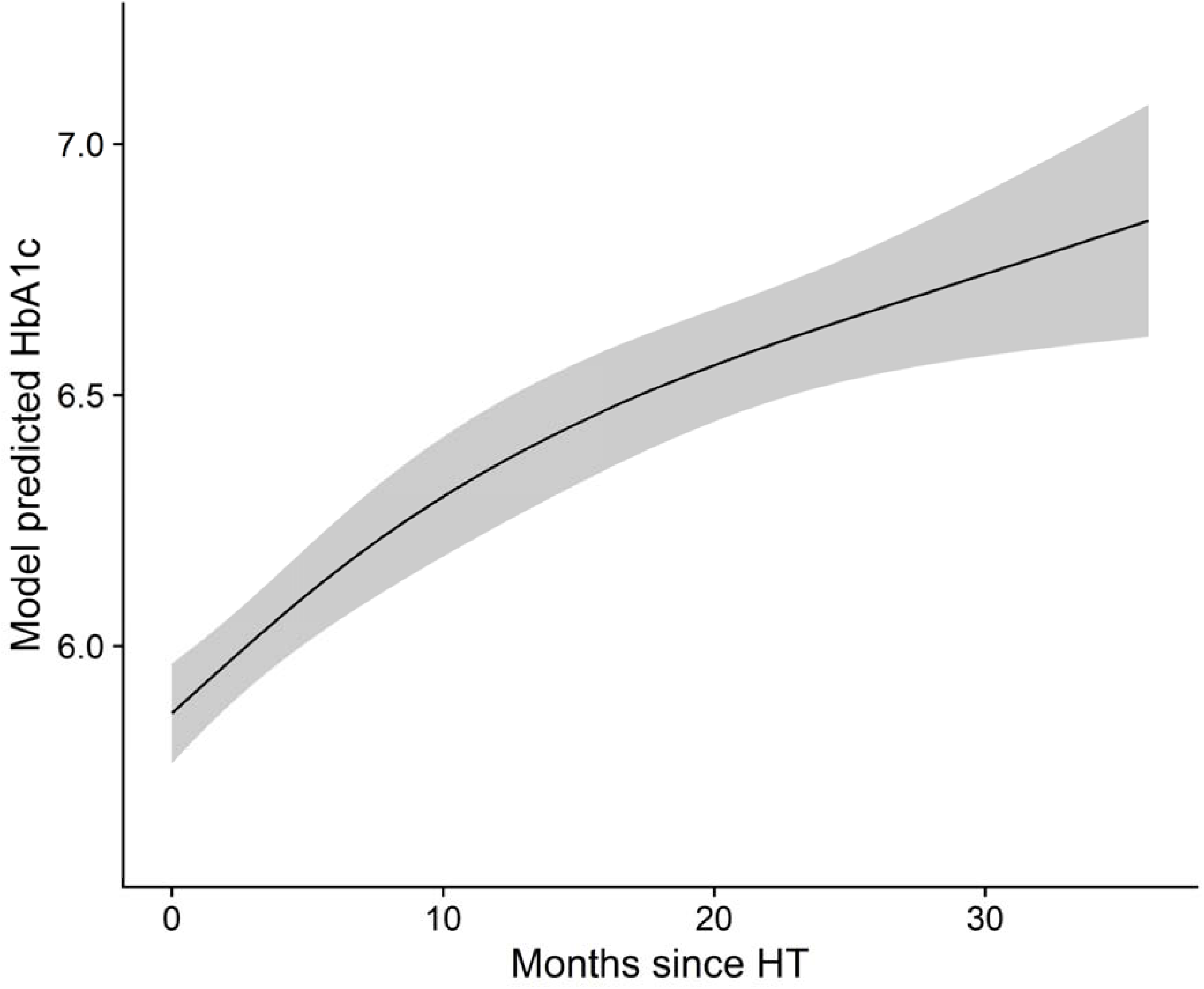
(**A**) Kaplan-Meier curve of proportion of adults free from DM2 following HT. **(B)** Kaplan-Meier curve of proportion of adults with HbA1c < 6.5% who develop HbA1c ≥6.5% following HT. **(C)** GLS modeling of HbA1c trajectory in the 36 months following HT.

**Table 2:**
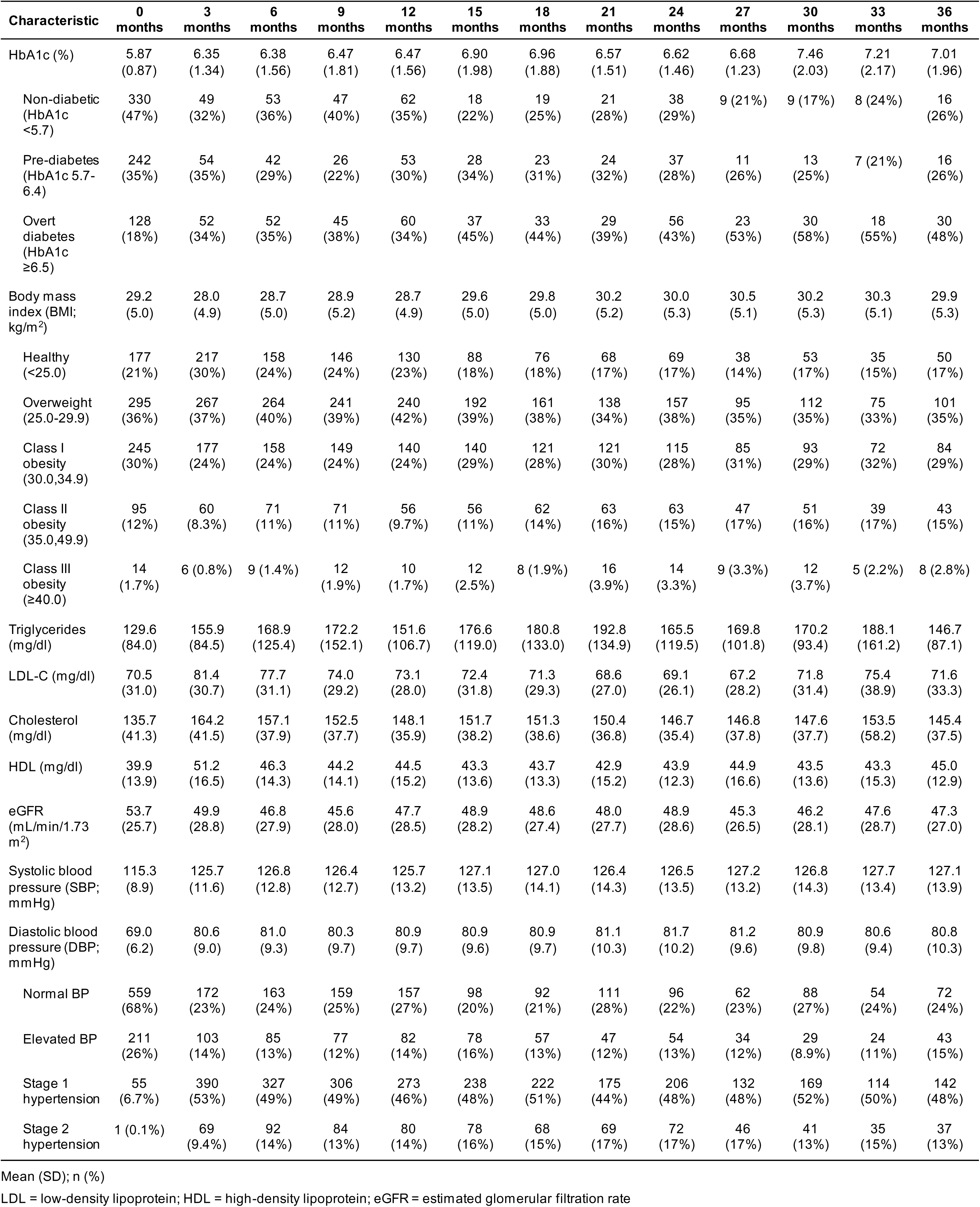
Post-transplant CKM characteristics of adult heart transplant recipients.

### Trends in body mass index following heart transplantation in adults

Among 826 adults with available BMI data at HT, mean BMI was as 29.2 ± 5.0 kg/m^2^. Of these, 21.4% had underweight or healthy weight, 35.7% had overweight, 29.7% had class I obesity, 11.5% had class II obesity, and 1.7% had class III obesity (**Table 2**). The prevalence of post-HT underweight/healthy weight, overweight, and obesity is depicted in **Figure 2A**. Among 178 adults with BMI < 25 kg/m^2^ at time of HT, the cumulative incidences of developing a BMI ≥ 25 kg/m^2^ within 6-months and 1-year post-HT were 56.6% and 66.9%, respectively (**Figure 2B**). The IR of overweight or obesity was 77.3 per 100 person-years (95% CI 63.8-90.7 per 100 person-years). GLS modeling demonstrated time after HT was a significant predictor of BMI trajectory (P_chunk_ = 0.001; **Figure 2C**), showing significant linear (β = –0.017, P-value <0.001) and non-linear (β = 0.052, P-value = 0.001) time effects. After HT, there was an initial predicted decline in BMI in all adults, followed by a steady increase in BMI thereafter. Among 168 individuals started on a GLP1ra (semaglutide, exenatide, dulaglutide or lixisenatide) after HT, there was an overall significant change in BMI in the 12 months following prescription (P_chunk_ = 0.006; **Figure 2D**), with a predominantly linear decrease (linear β = –0.080, P-value = 0.10).

**Figure 2.**
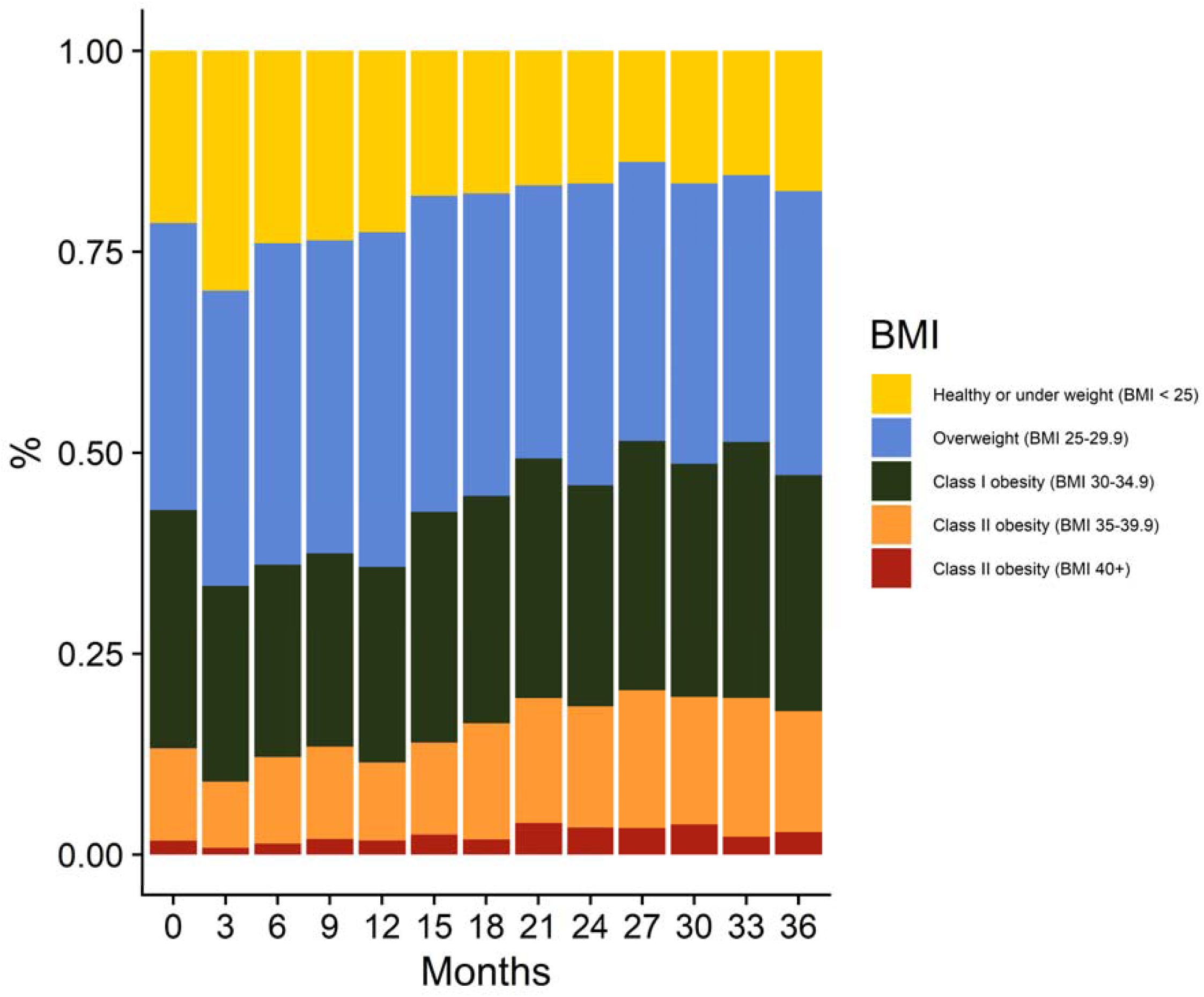
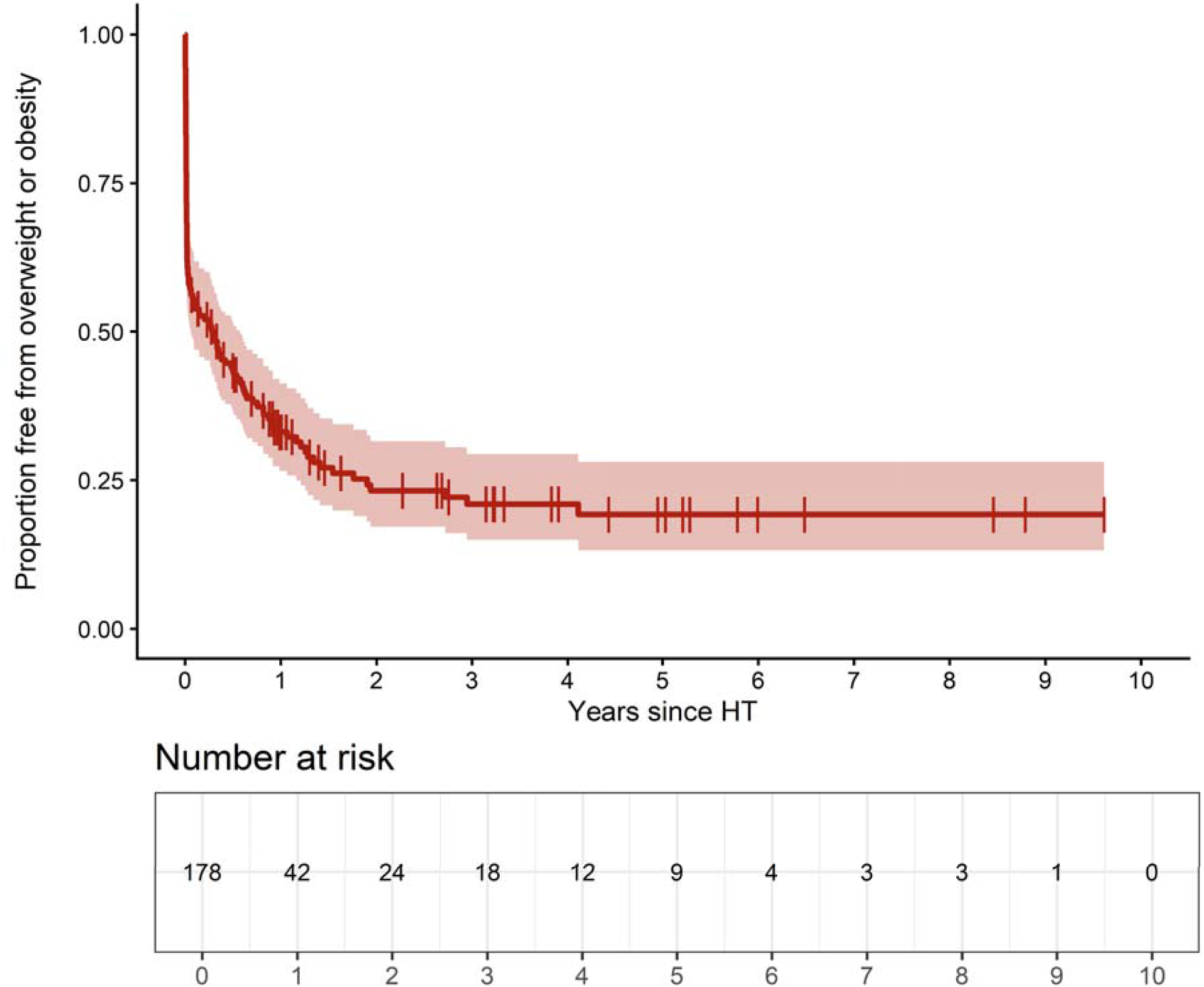
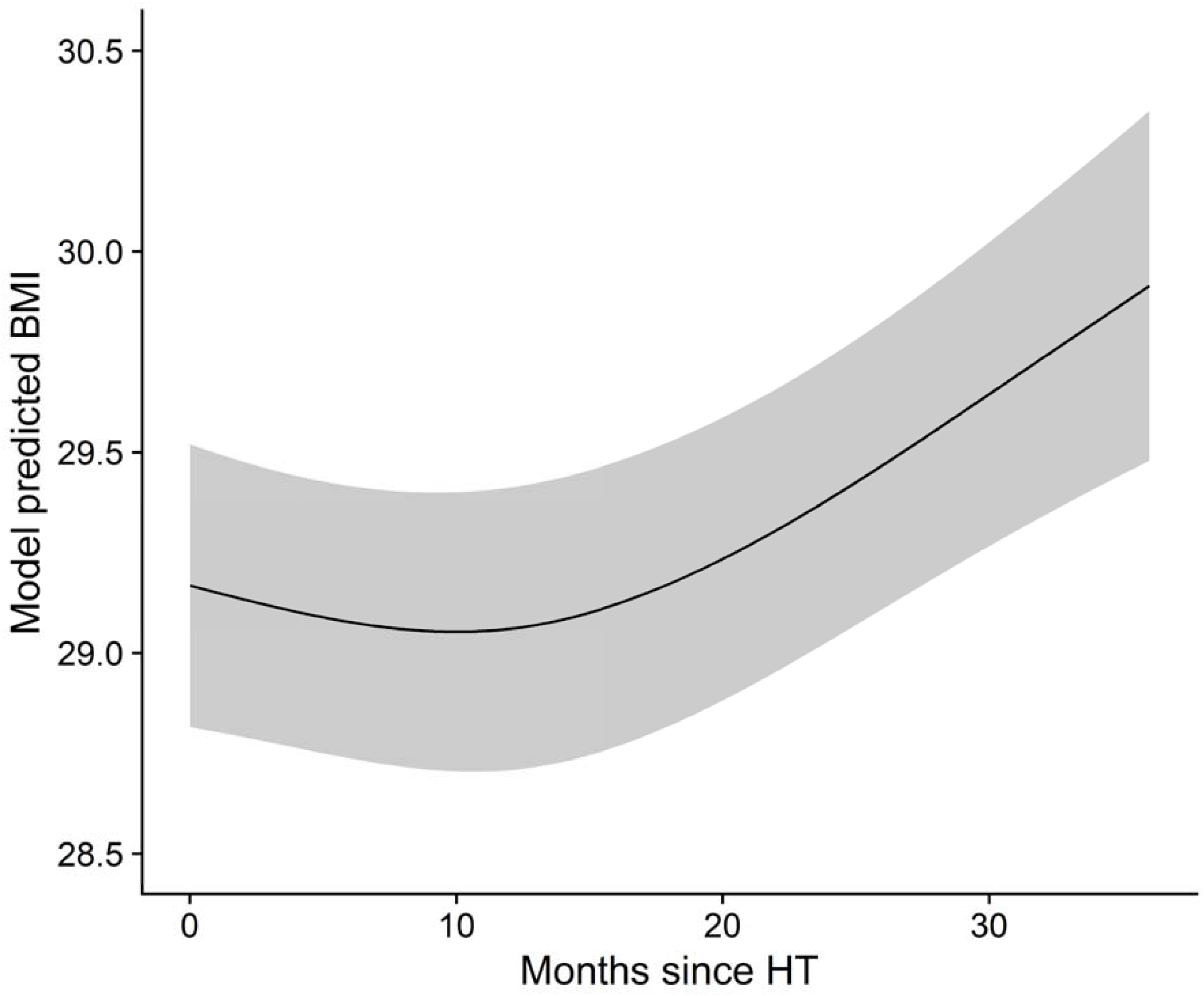
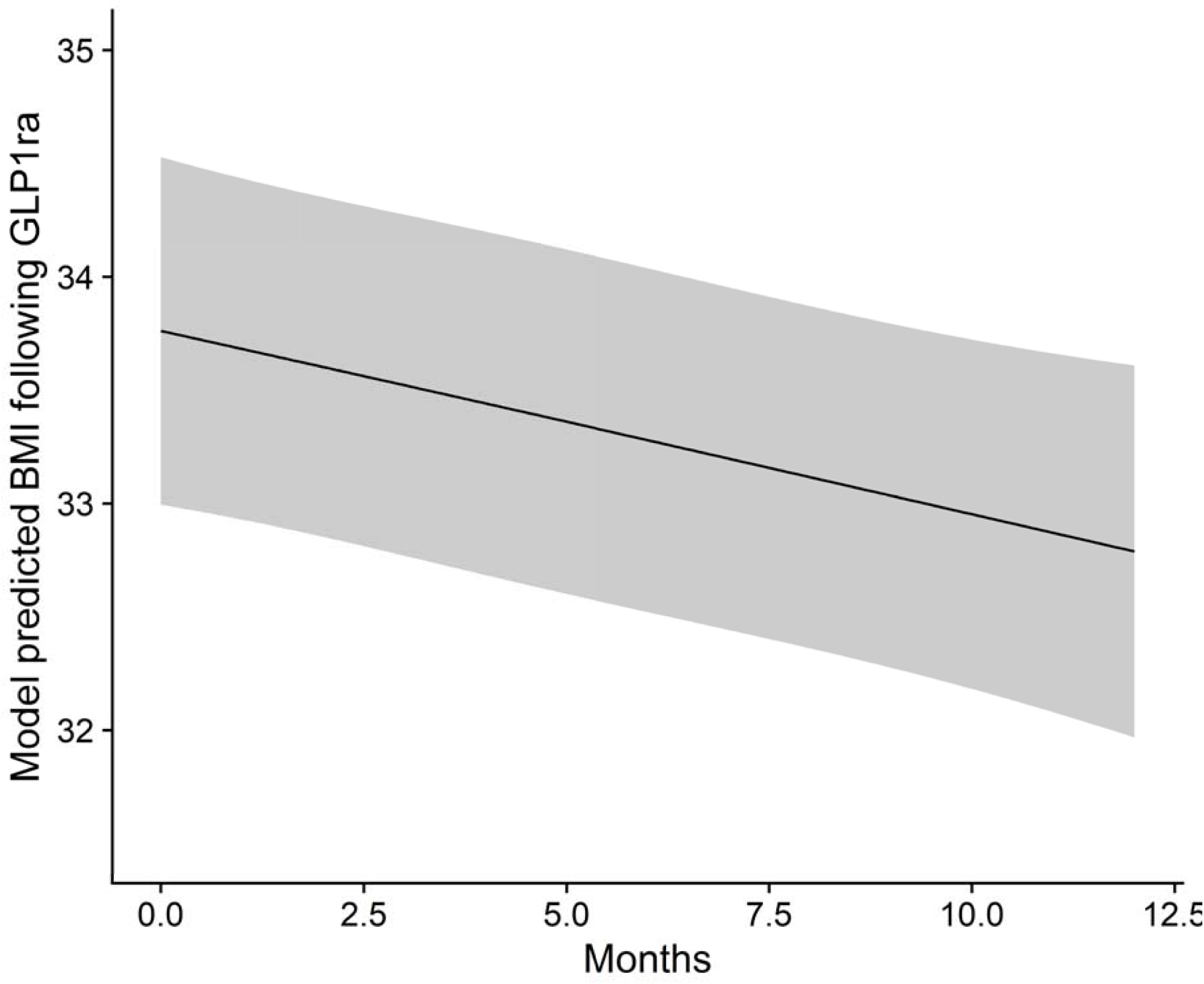
(**A**) Distribution of healthy or normal weight, overweight, and obesity in adults at the time of HT and in the 36 months following HT. **(B)** Kaplan-Meier curve of proportion of adults free from overweight or obesity following HT. **(C)** GLS modeling of BMI trajectory in the 36 months following HT. **(D)** GLS modeling of changes in BMI trajectory in the 12 months following initiation of GLP1ra therapy.

### Trends in lipid profiles following heart transplantation in adults

Most adults (N = 653) had a diagnosis of dyslipidemia prior to HT. At the time of HT, the mean triglyceride level was 129.6 ± 84.0 mg/dl, mean LDL-C level was 70.5 ± 31.0 mg/dl, mean total cholesterol level was 135.7 ± 41.3 mg/dl, and mean HDL level was 39.9 ± 13.9 mg/dl. LDL-C, total cholesterol, and HDL levels all uptrended after transplant, peaking at 3 months at 81.4 ± 30.7 mg/dl, 164.2 ± 41.5 mg/dl and 51.2 ± 16.5 mg/dl, respectively (**Table 2**). Triglyceride levels also uptrended steadily but didn’t peak until 21 months post-HT (192.8 ± 134.9 mg/dl). Among 207 adults without dyslipidemia prior to HT, the IR of dyslipidemia was 139.1 per 100 person-years (95% CI 118.7-159.5 per 100 person-years), with cumulative incidence of 77.7% of individuals developing dyslipidemia by 6 months post-HT (**Figure 3A**). In 572 adults with triglycerides <200 mg/dl at time of HT, 37.5% developed levels ≥200 mg/dl by 1-year post HT. Similarly, in 470 adults with LDL-C <100 mg/dl at the time of HT, 31.1% developed levels ≥100 mg/dl by 1-year post HT. This translated into an IR of 29.4 per 100 person-years (95% CI 26.0-32.8 per 100 person-years) for worsening triglyceridemia post-HT and 21.7 per 100 person-years (95% CI 18.6-24.8 per 100 person-years) for worsening LDL-C control post-HT. GLS modeling demonstrated significant linear and nonlinear time effect after HT to increasing triglycerides (P_chunk_ <0.001) and cholesterol (P_chunk_ = 0.007), with a predominantly linear time effect on LDL-C (P_chunk_ = 0.029). In our modeling, triglycerides were predicted to peak between 15 and 18 months post-HT (linear β = 2.782, P-value <0.001; nonlinear β = –3.231, P-value <0.001; **Figure 3B**) while total cholesterol levels were predicted to peak between 12 and 15 months post-HT (linear β = 0.560, P-value = 0.007; nonlinear β = – 0.866, P-value = 0.002; **Figure 3C**). On the other hand, LDL-C levels were predicted to steadily decline following HT (linear β = –0.172, P-value = 0.029; nonlinear β = 0.035, P-value = 0.871; **Figure 3D**). There were no significant changes in HDL levels post-HT (P_chunk_ = 0.227).

**Figure 3.**
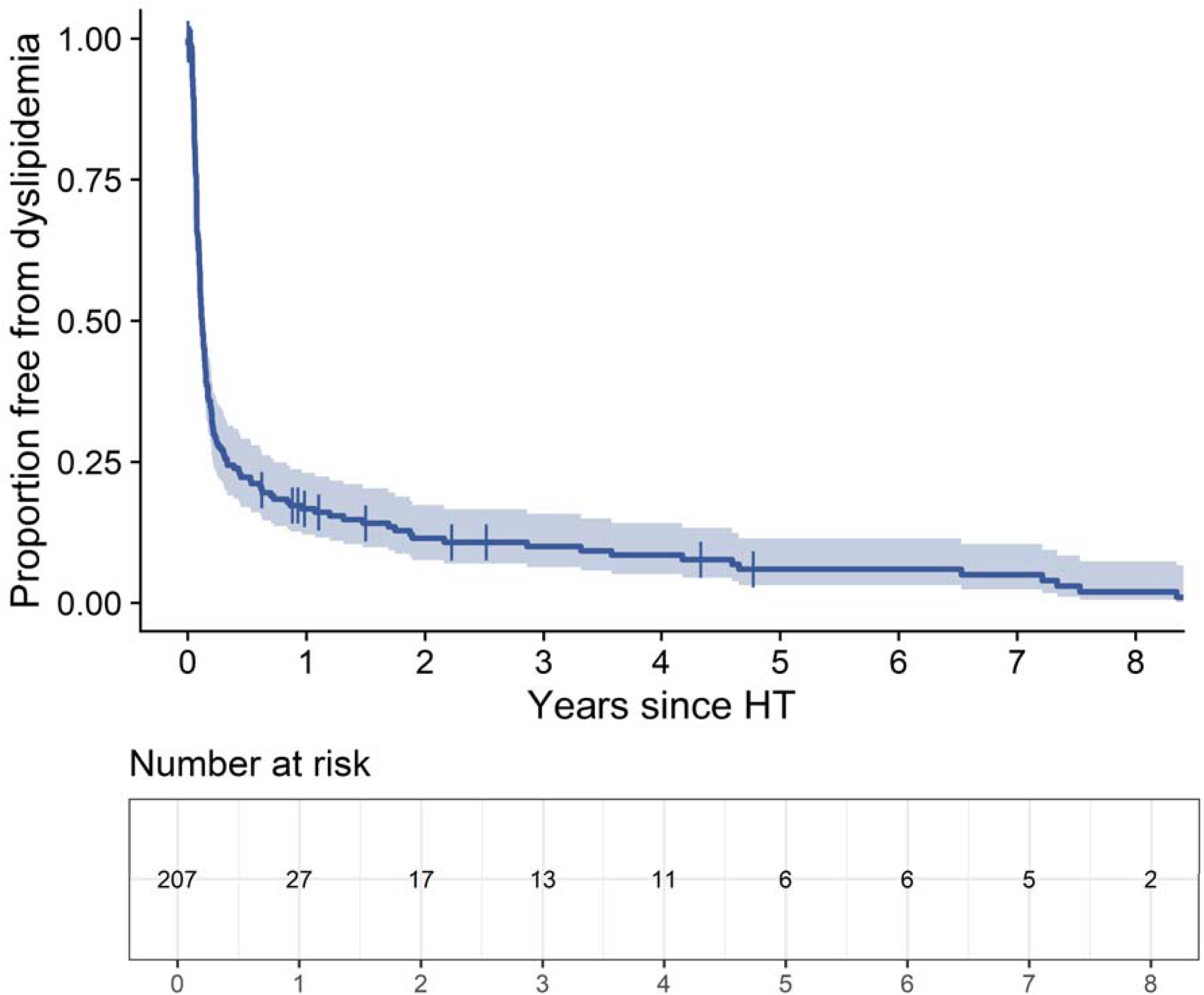
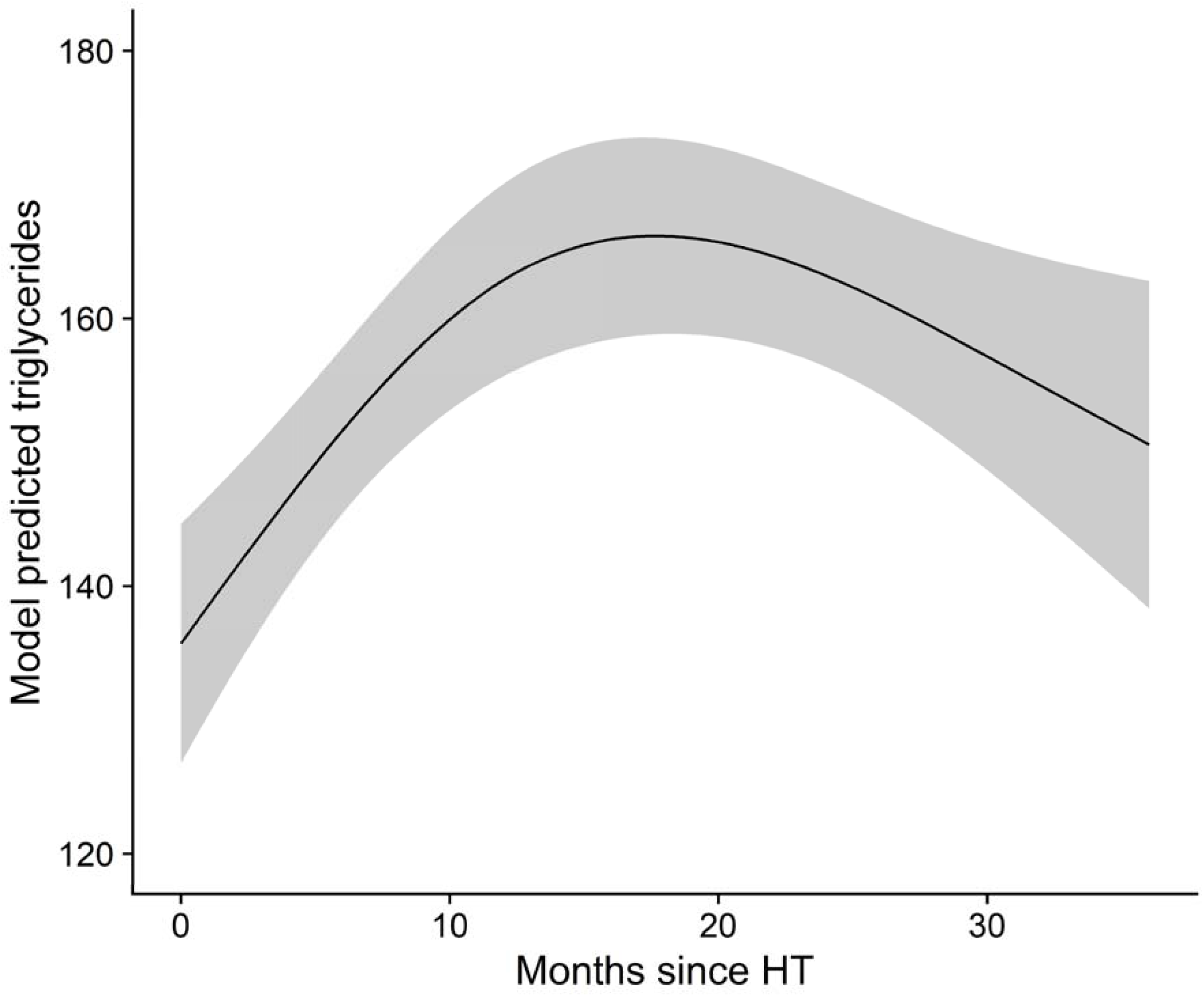
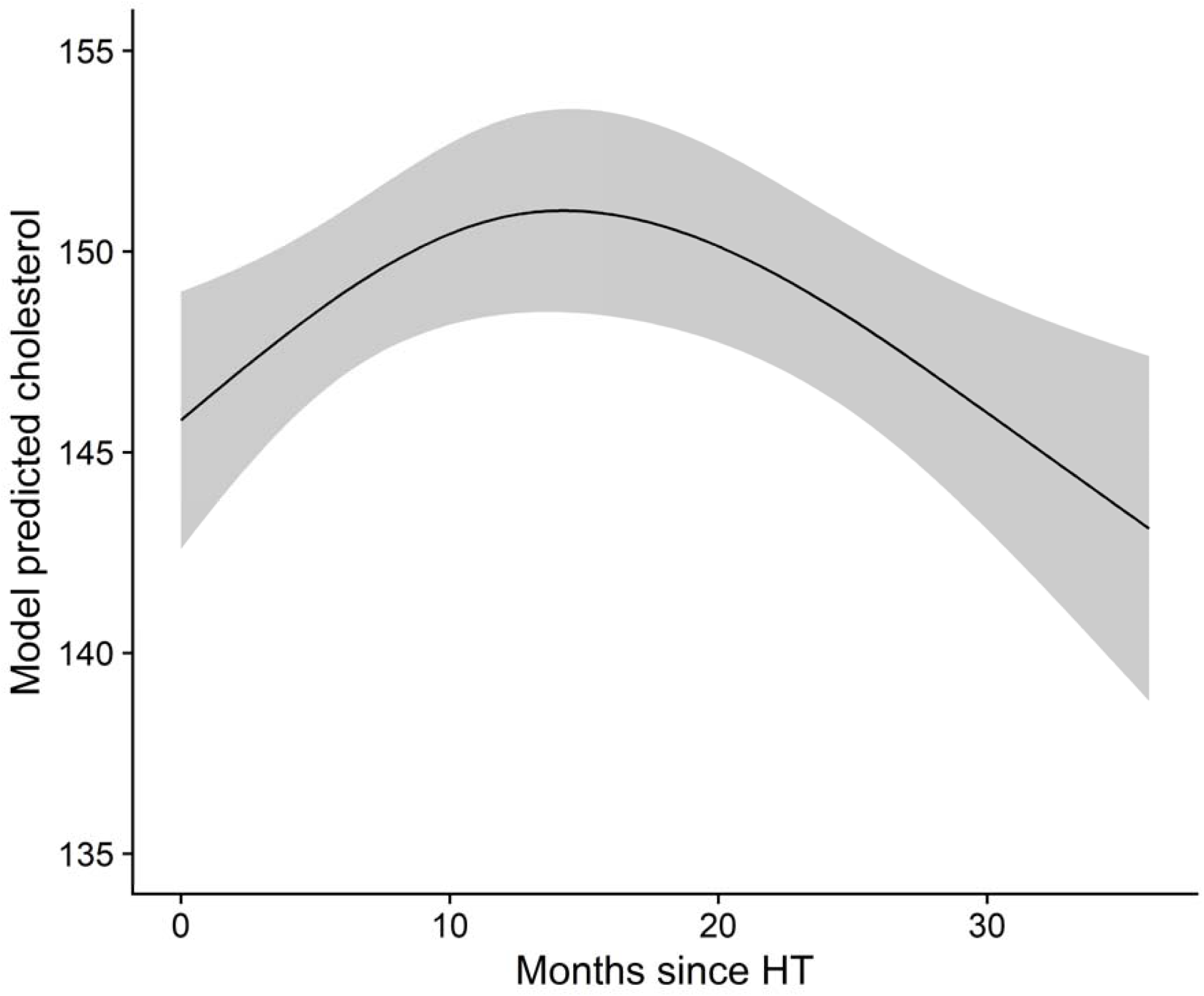
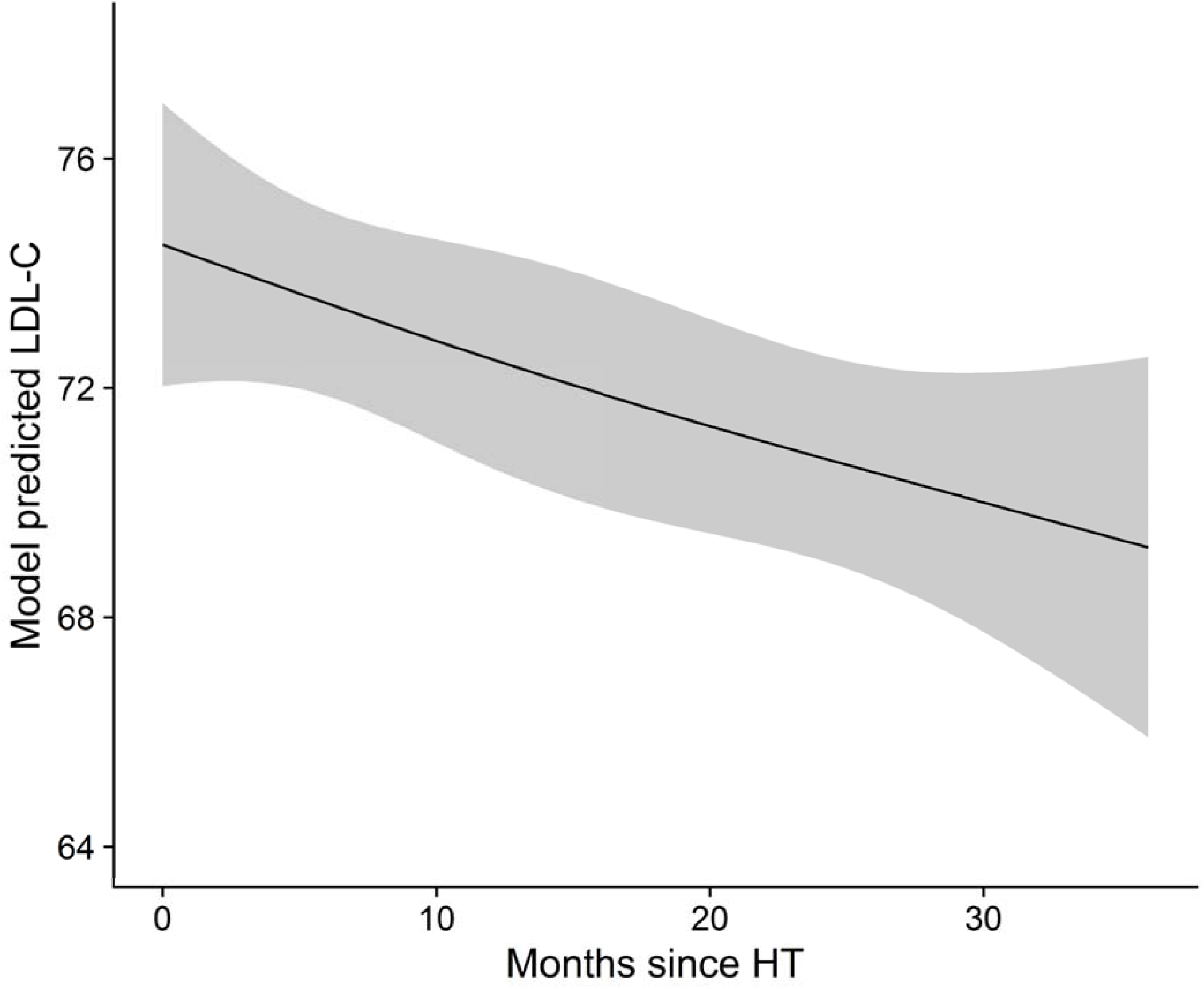
(**A**) Kaplan-Meier curve of proportion of adults free from dyslipidemia following HT. **(B)** GLS modeling of triglyceride trajectory in the 36 months following HT. **(C)** GLS modeling of total cholesterol trajectory in the 36 months following HT. **(D)** GLS modeling of LDL-C trajectory in the 36 months following HT.

### Trends in hypertension after heart transplantation in adults

Prior to HT, 67.7% of adults had normal BP, 25.5% had elevated BP, 6.7% had stage 1 hypertension, and 0.1% had stage 2 hypertension (**Table 2**). Mean pre-HT SBP was 115.3 ± 8.9 mmHg and mean pre-HT DBP was 69.0 ± 6.2 mmHg (**Figure 4A**). Among adults with normal or elevated BP measurements preceding HT, the cumulative incidence of developing stage 1 or more hypertension by 6 months post-HT was 98.1%. GLS models revealed significant linear and nonlinear contributions of time from HT (P_chunk_ <0.001) to increasing SBP and DBP (**Figure 4B and 4C**). Both SBP (linear β = 0.945, nonlinear β = –0.980; P-value <0.001 for both) and DBP (linear β = 1.072, nonlinear β = –1.141; P-value <0.001 for both) increased post-HT and were predicted to peak between 18 and 21 months.

**Figure 4.**
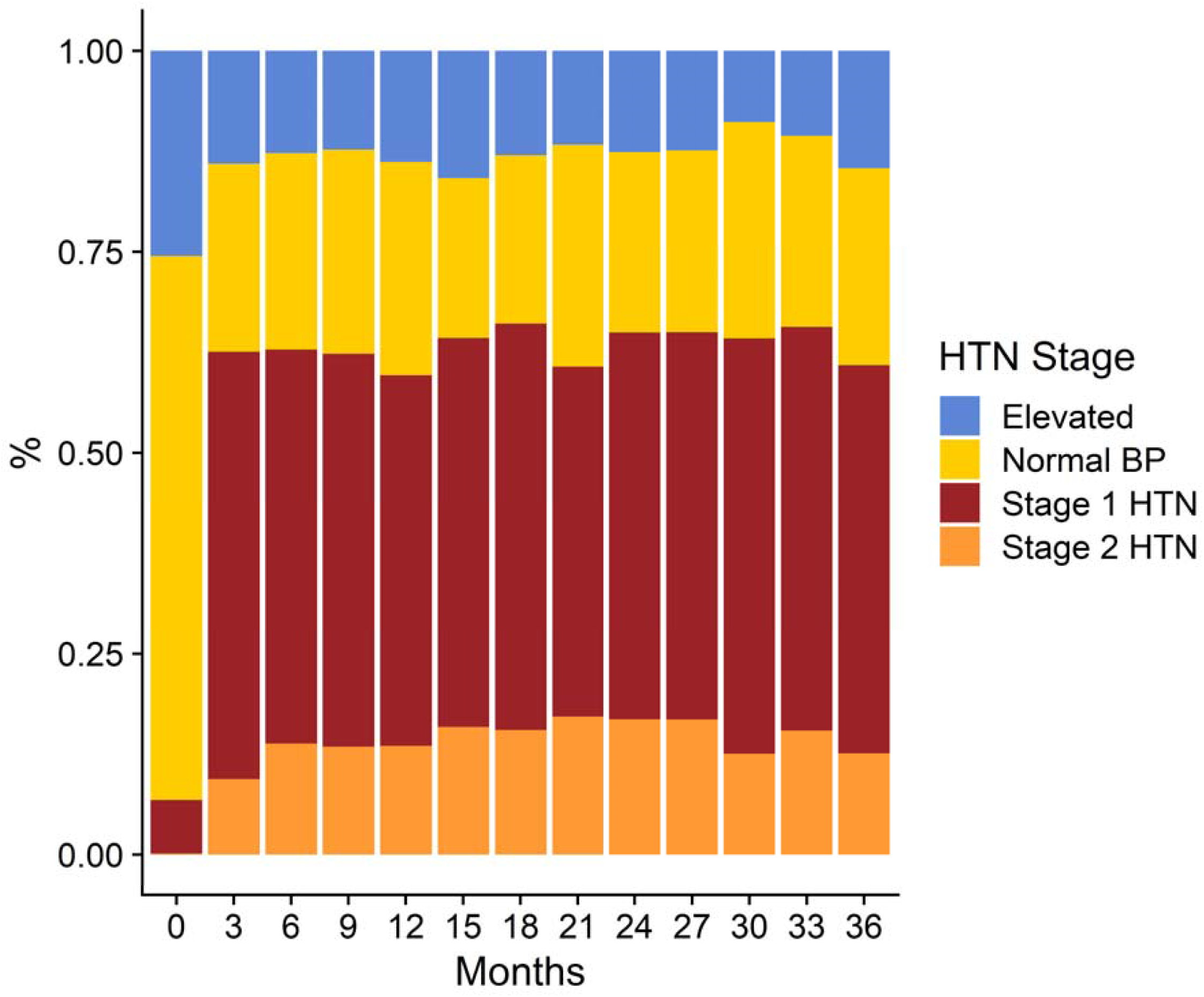
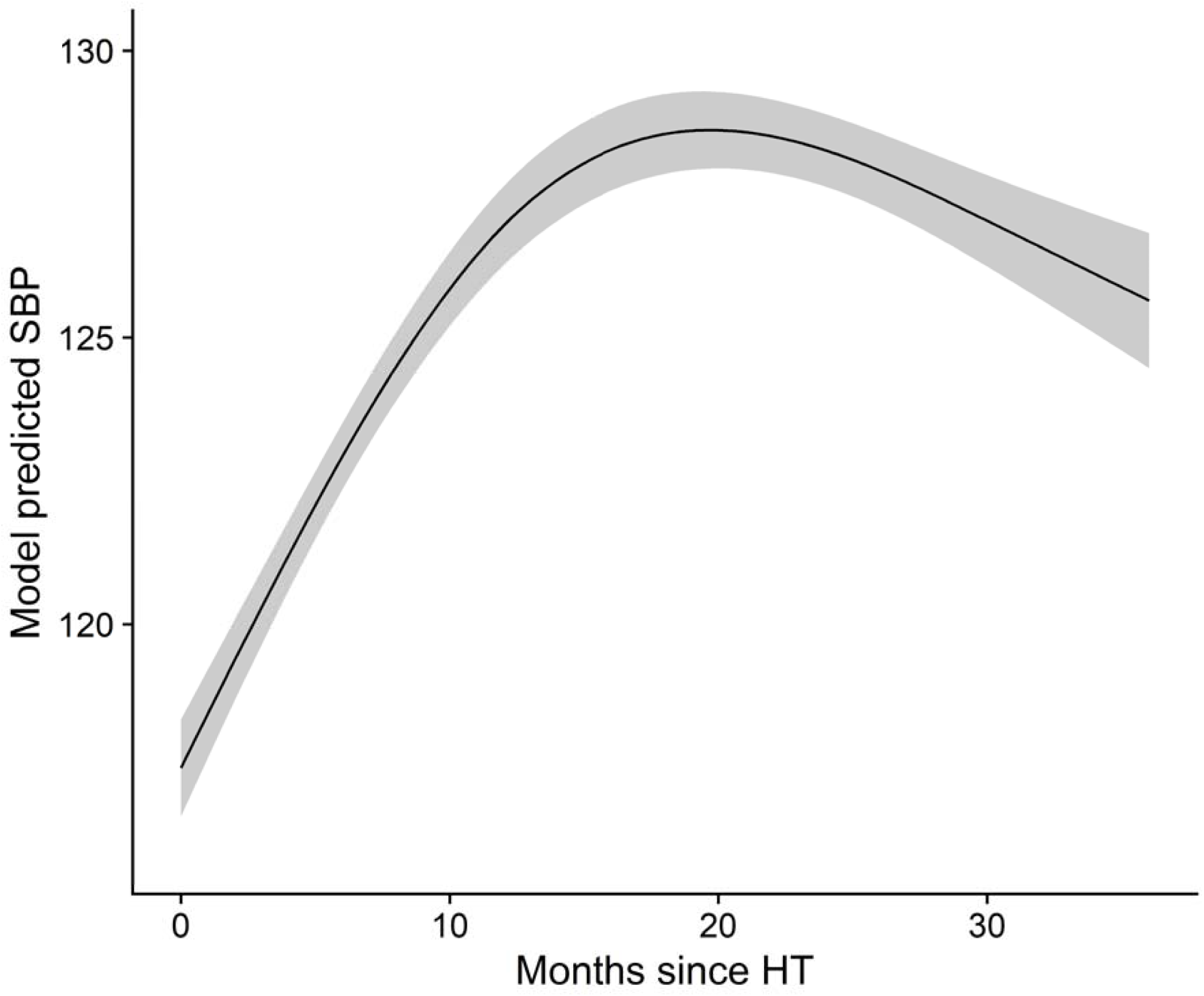
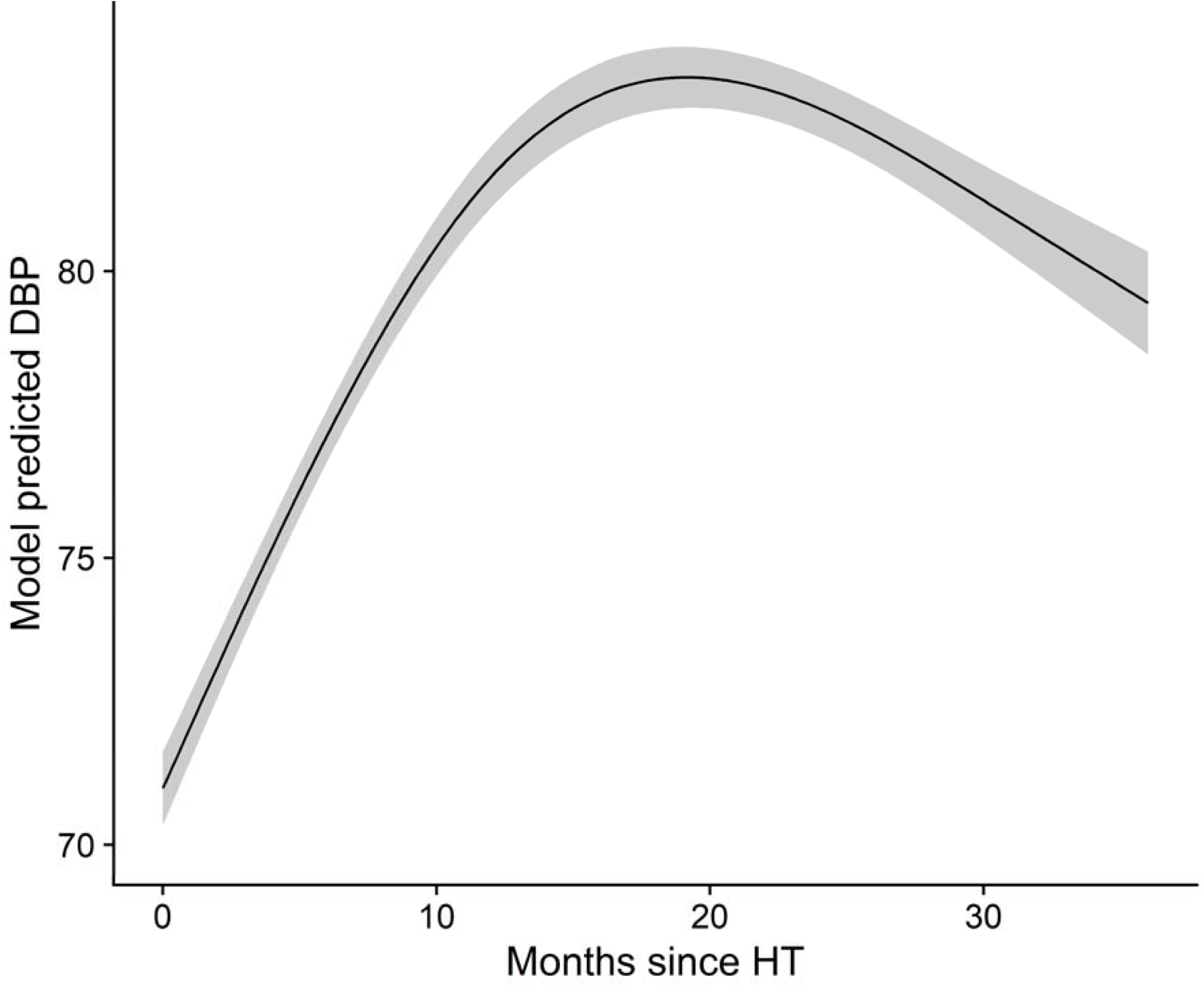
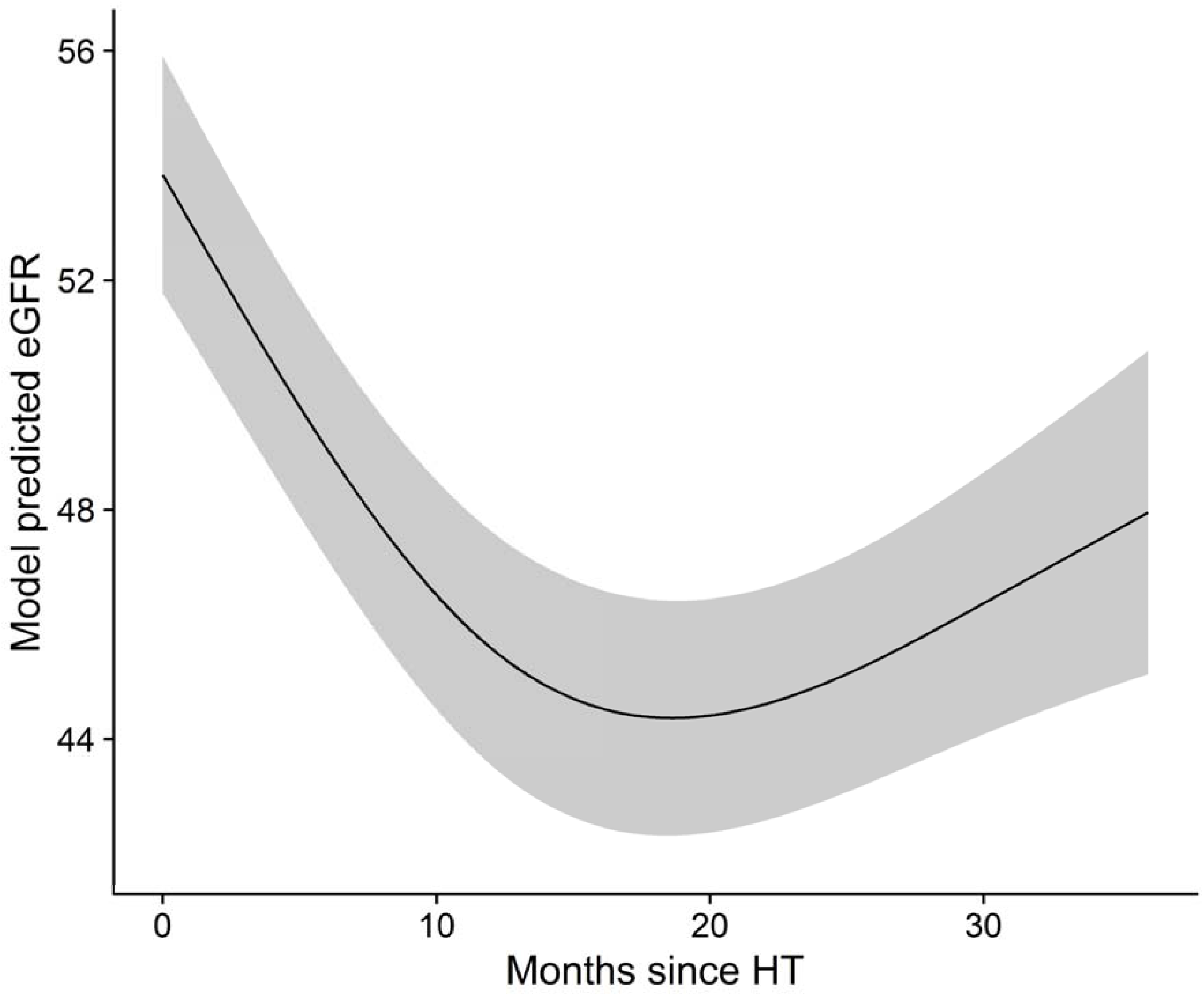
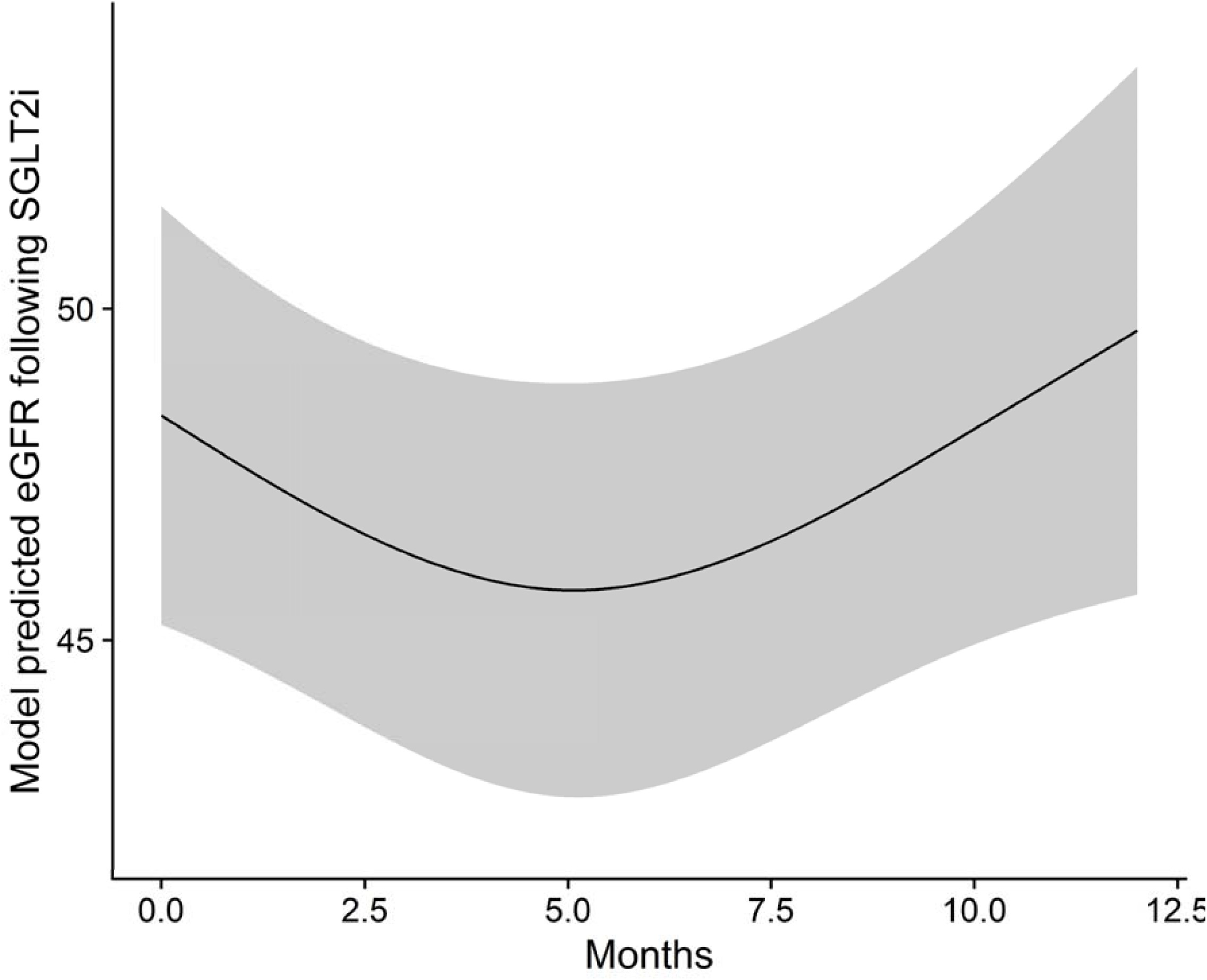
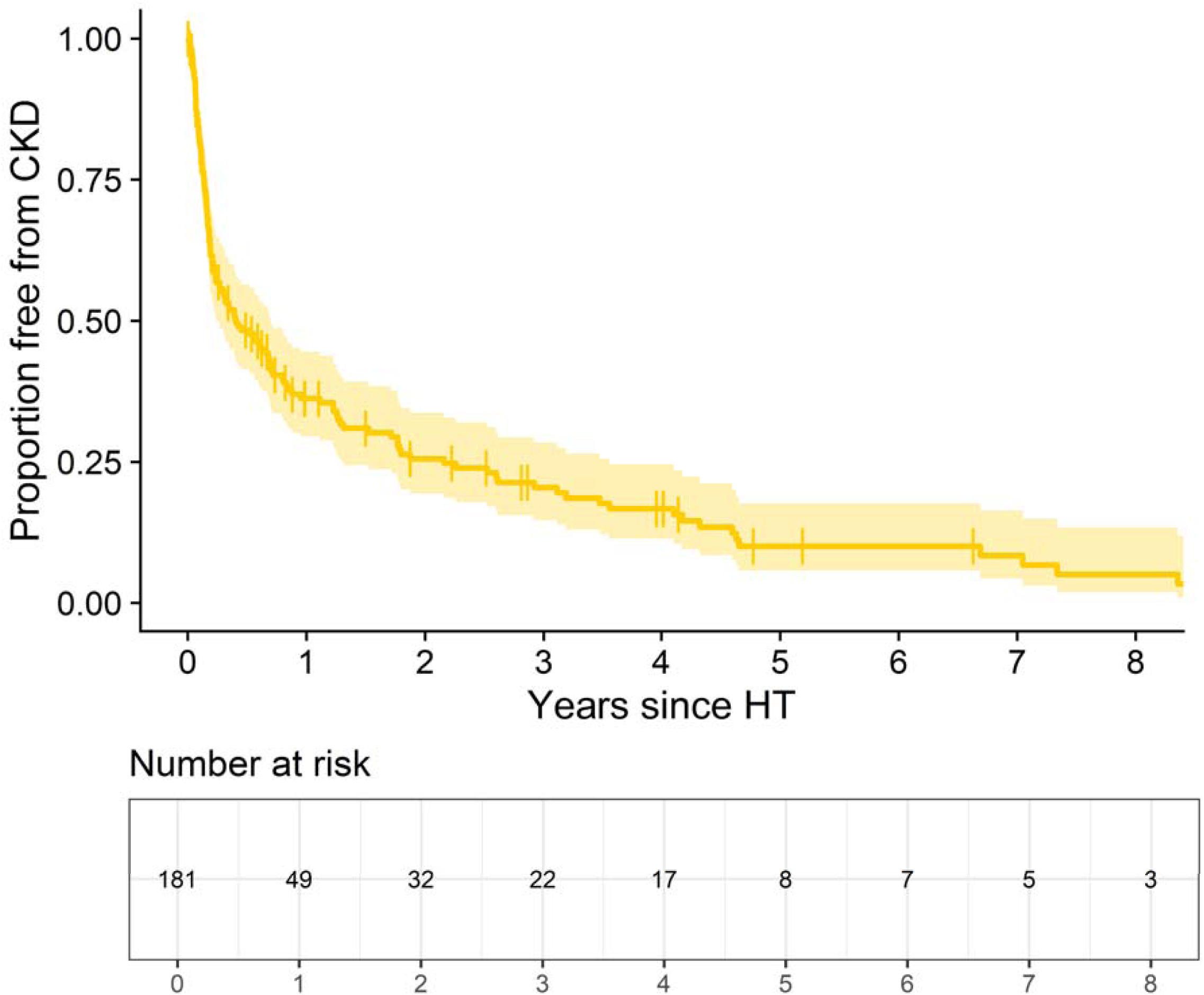
(**A**) Distribution of BP categories at the time of HT and in the 36 months following HT. **(B)** GLS modeling of systolic BP in the 36 months following HT. **(C)** GLS modeling of diastolic BP in the 36 months following HT. **(D)** GLS modeling of eGFR in the 36 months following HT. **(E)** Kaplan-Meier curve of proportion of adults free from CKD following HT. **(F)** GLS modeling of changes in eGFR trajectory in the 12 months following initiation of SGLT2 inhibitor therapy.

### Trends in renal function after heart transplantation in adults

After excluding individuals who required dialysis pre-HT or underwent multiorgan transplant, mean eGFR at the time of HT was 53.7 ± 25.7 mL/min/1.73 m^2^ and steadily decreased thereafter, reaching a nadir of 45.3 ± 26.5 mL/min/1.73 m^2^ at 27 months post-HT (**Table 2**). Among 583 adults with eGFR ≥45 mL/min/1.73 m^2^ pre-HT, the cumulative incidence of experiencing worsening eGFR to <45 mL/min/1.73 m^2^ within 6-months post-HT was 81.8%, translating to an IR of 186.8 per 100 person-years (95% CI 170.8-202.8 per 100 person-years; **Figure 4D**). Among 181 adults without CKD pre-HT, 51.7% and 63.8% developed CKD within 6 months and 1-year post-HT, respectively (**Figure 4E**). This translated to an IR of a new CKD diagnosis of 69.7 per 100 person-years (95% CI 58.2-81.3 per 100 person-years). GLS modeling demonstrated significant linear and nonlinear time effects from HT (P_chunk_ <0.001) to declining eGFR. In 242 individuals who were prescribed SGLT2i post-HT, there were significant linear and nonlinear time effects on eGFR during the ensuing 12 months (linear β = –0.781, P-value = 0.0384 nonlinear β = 1.014, P-value = 0.016; **Figure 4F**).

### Trends in pediatric cardiometabolic disease following transplantation

In children, the IR of post-HT DM2 was 2.8 per 100 person-years (95% CI 1.0-4.6 per 100 person-years). Only 14 children had HbA1c measured prior to HT, among whom mean HbA1c was 5.7 ± 0.5% (**Table 3**). The mean BMI z-score among 74 children who had BMI measured prior to HT was 0.41 ± 1.36 at time of HT, with 66.2% of children having normal weight, 14.9% having overweight, and 18.9% having obesity (**Figure 5A**). Of 49 children who were underweight or normal weight, the cumulative incidence of developing overweight or obesity (BMI z-score ≥1.04) by 6-months post-HT was 43% (**Figure 5B**), which translated to an IR of 26.9 per 100 person-years (95% CI 16.1-37.7 per 100 person-years). The IR of dyslipidemia was 5.5 per 100 person-years (95% CI 2.7-8.2 per 100 person-years; **Figure 5C**). Mean LDL-C among children (N = 13) was 69.9 ± 60.4 mg/dl preceding HT, initially peaking at 79.5 ± 21.7 mg/dl at 3 months post-HT followed by a downtrend and another peak of 80.0 ± 13.0 mg/dl at 27 months post-HT. Similar trends were seen with total cholesterol (113.0 ± 40.5 mg/dl pre-HT, 154.7 ± 31.5 mg/dl at 3 months post-HT, and 177.1 ± 33.9 mg/dl at 33 months post-HT) and triglycerides (144.6 ± 86.9 mg/dl pre-HT, 176.7 ± 109.8 mg/dl at 3 months post-HT, and 491.9 ± 294.4 mg/dl at 33 months post-HT). Of 23 children with LDL-C <100 mg/dl preceding HT, the cumulative incidence of worsening LDL-C to ≥100 mg/dl by 1-year post-HT was 9.5%, translating to an IR of 12.7 per 100 person-years (95% CI 3.9-21.5 per 100 person-years) for worsening LDL-C control. Among 33 children with triglycerides <200 mg/dl prior to HT, the cumulative incidence of developing worsening triglycerides to ≥200 mg/dl by 1-year post-HT was 21.6%, translating to an IR for worsening hypertriglyceridemia of 13.1 per 100 person-years (95% CI 6.0-20.2 per 100 person-years). Only two children developed stage 1 or greater hypertension. In GLS models for BP, SBP (linear β = 0.413, P-value = 0.009; nonlinear β = –0.514, P-value = 0.003) and DBP (linear β = 0.271, P-value = 0.007; nonlinear β = –0.284, P-value = 0.016) significantly changed over time, predicted to peak at 15 to 21 months post-HT. In evaluation of renal function, the IR for CKD diagnosis was 3.6 per 100 person-years (95% CI 1.5-5.8 per 100 person-years; **Figure 5D**). None of the pediatric individuals included in the study were prescribed SGLT2i or GLP1ras following HT.

**Figure 5.**
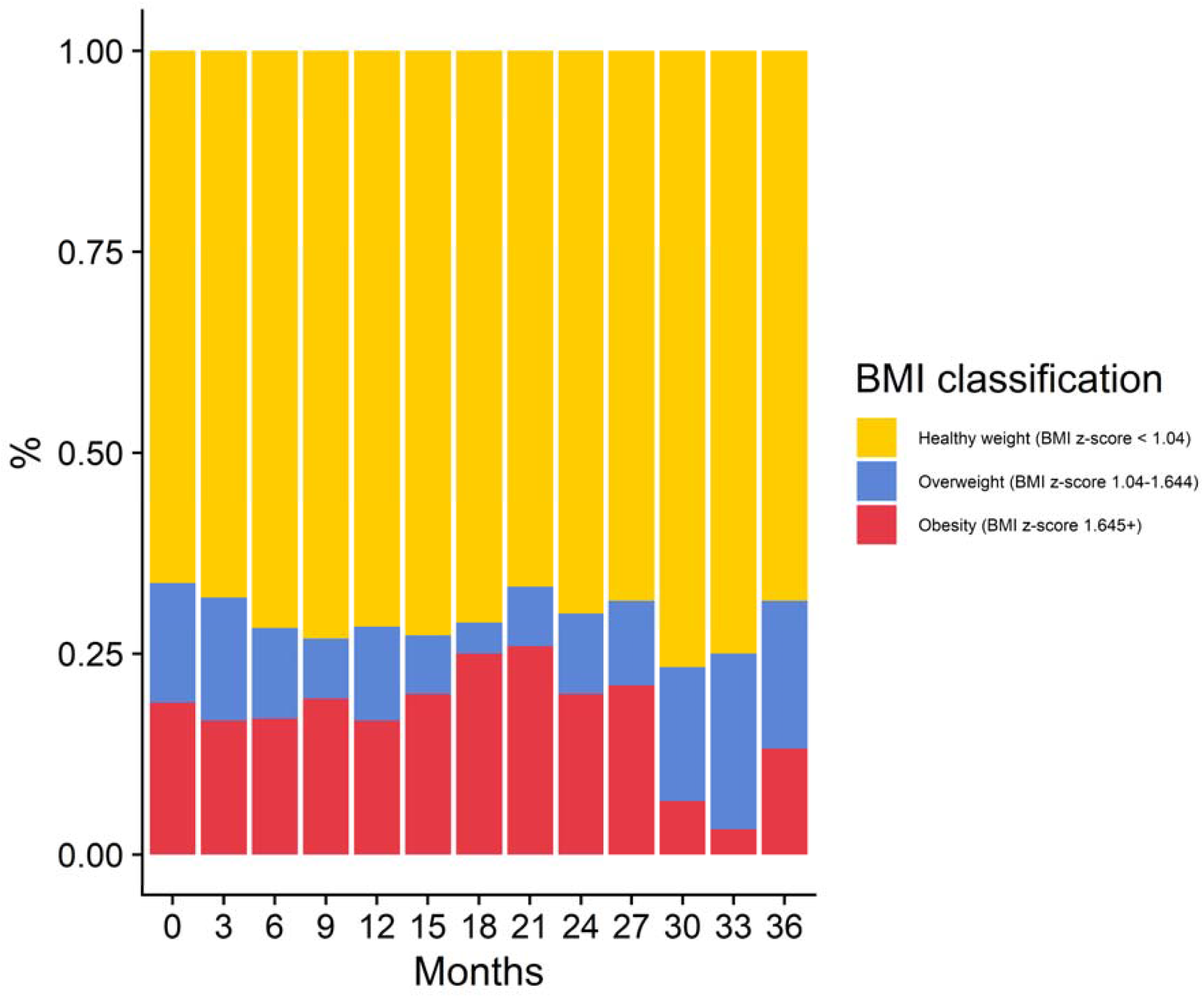
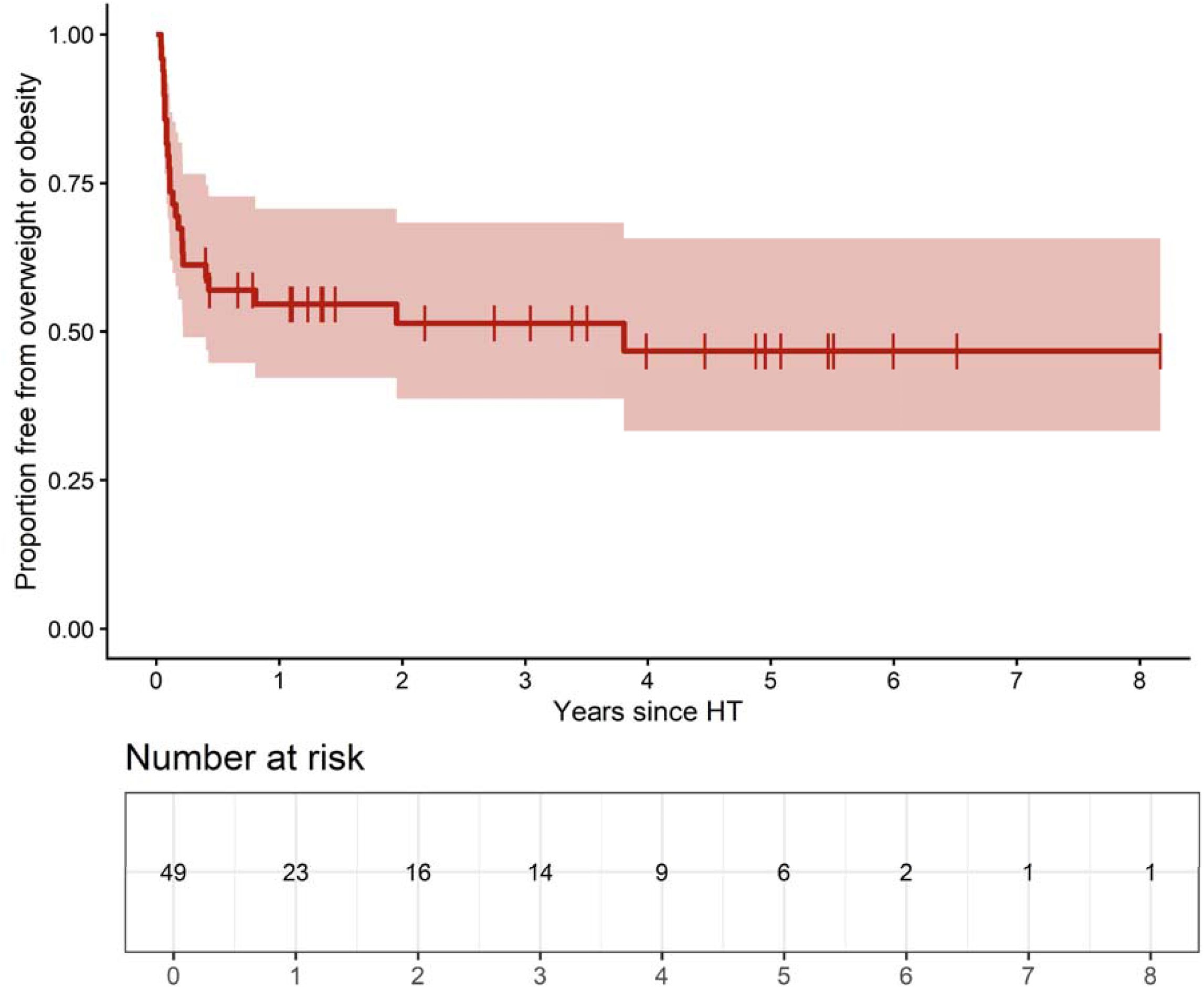
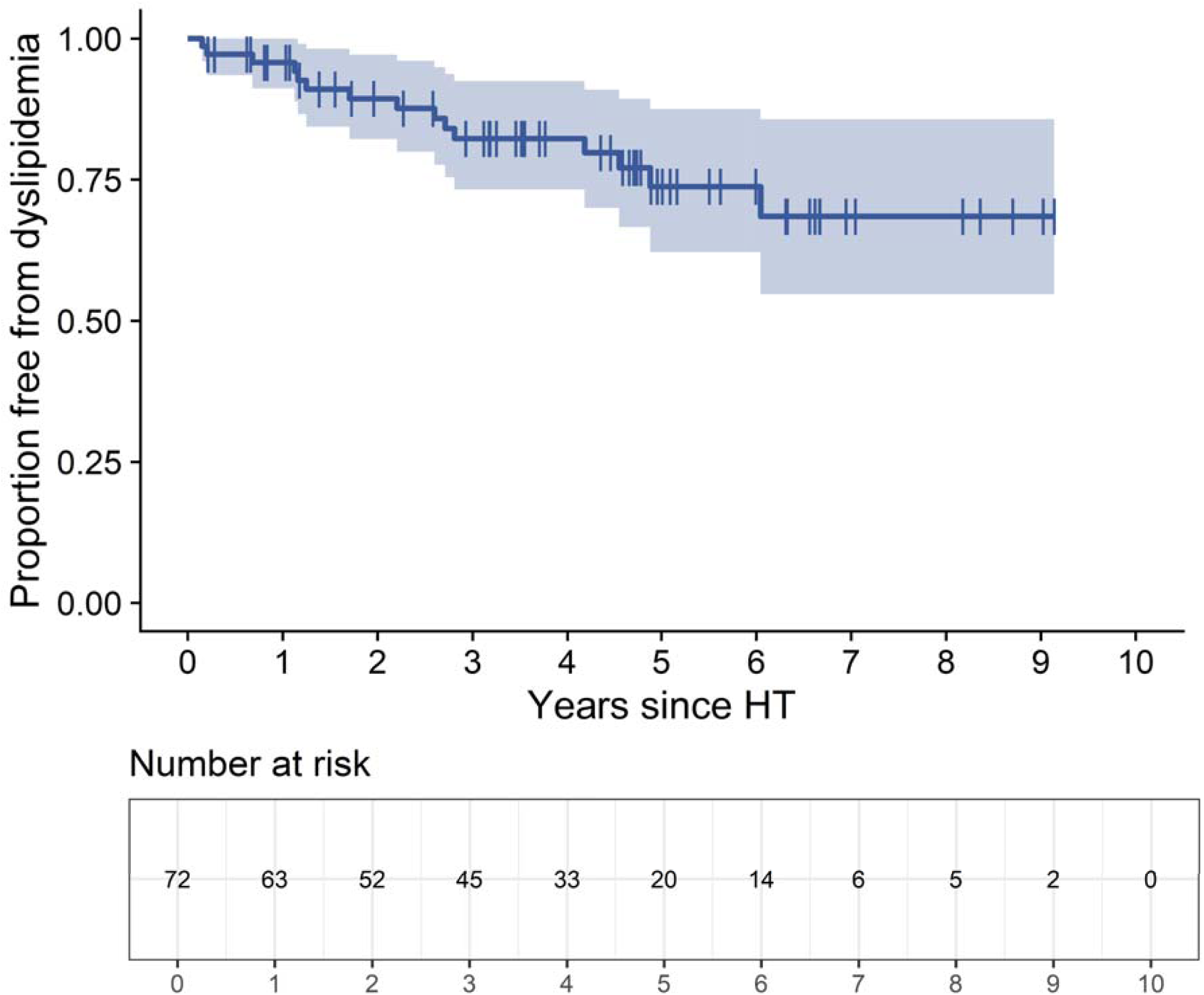
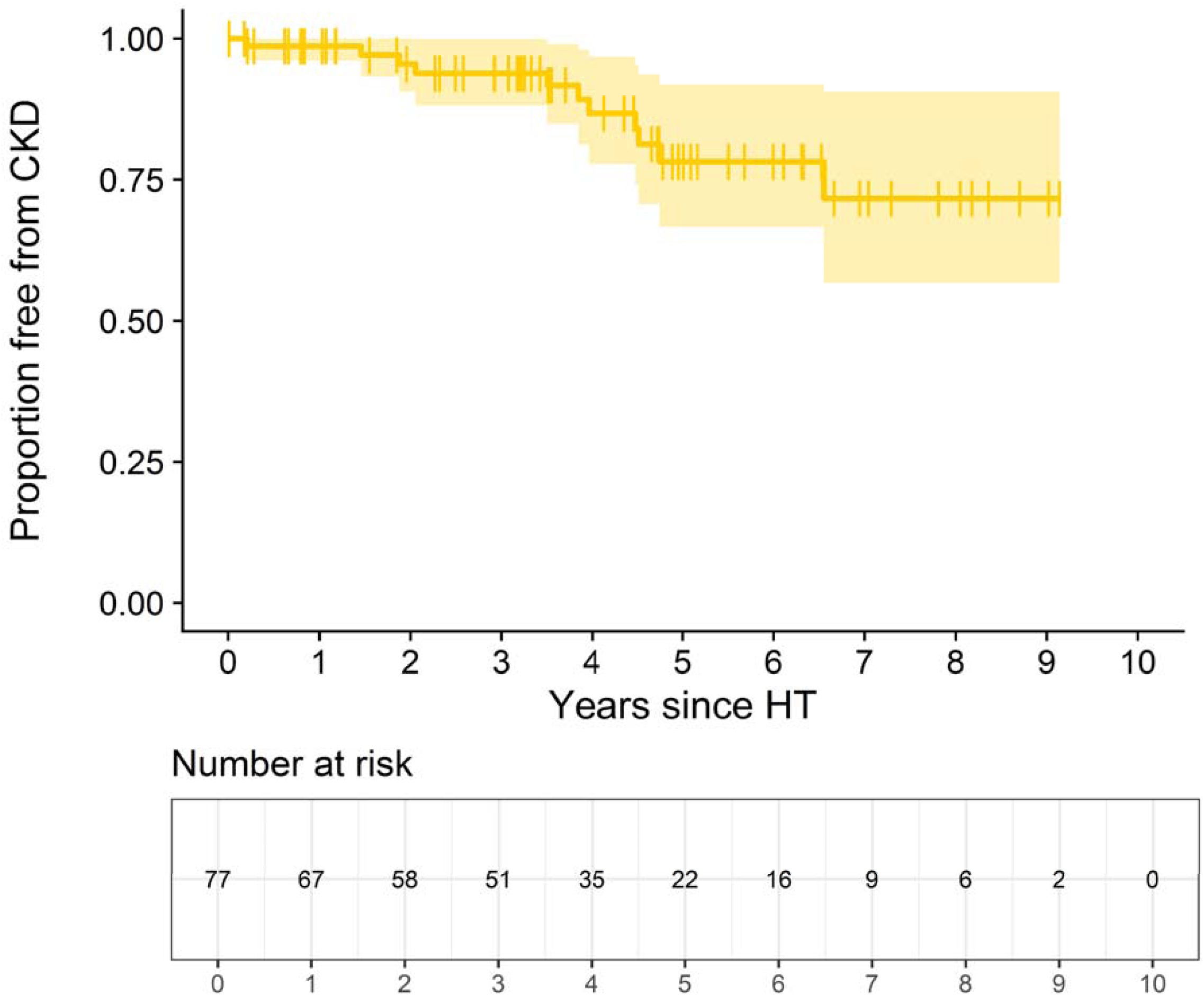
(**A**) Distribution of BMI z-scores in pediatric HT recipients in the 36 months following transplant. **(B)** Kaplan-Meier curve of proportion of children free from overweight or obesity following HT. **(C)** Kaplan-Meier curve of proportion of children free from dyslipidemia following HT. **(D)** Kaplan-Meier curve of proportion of children free from CKD following HT.

**Table 3:**
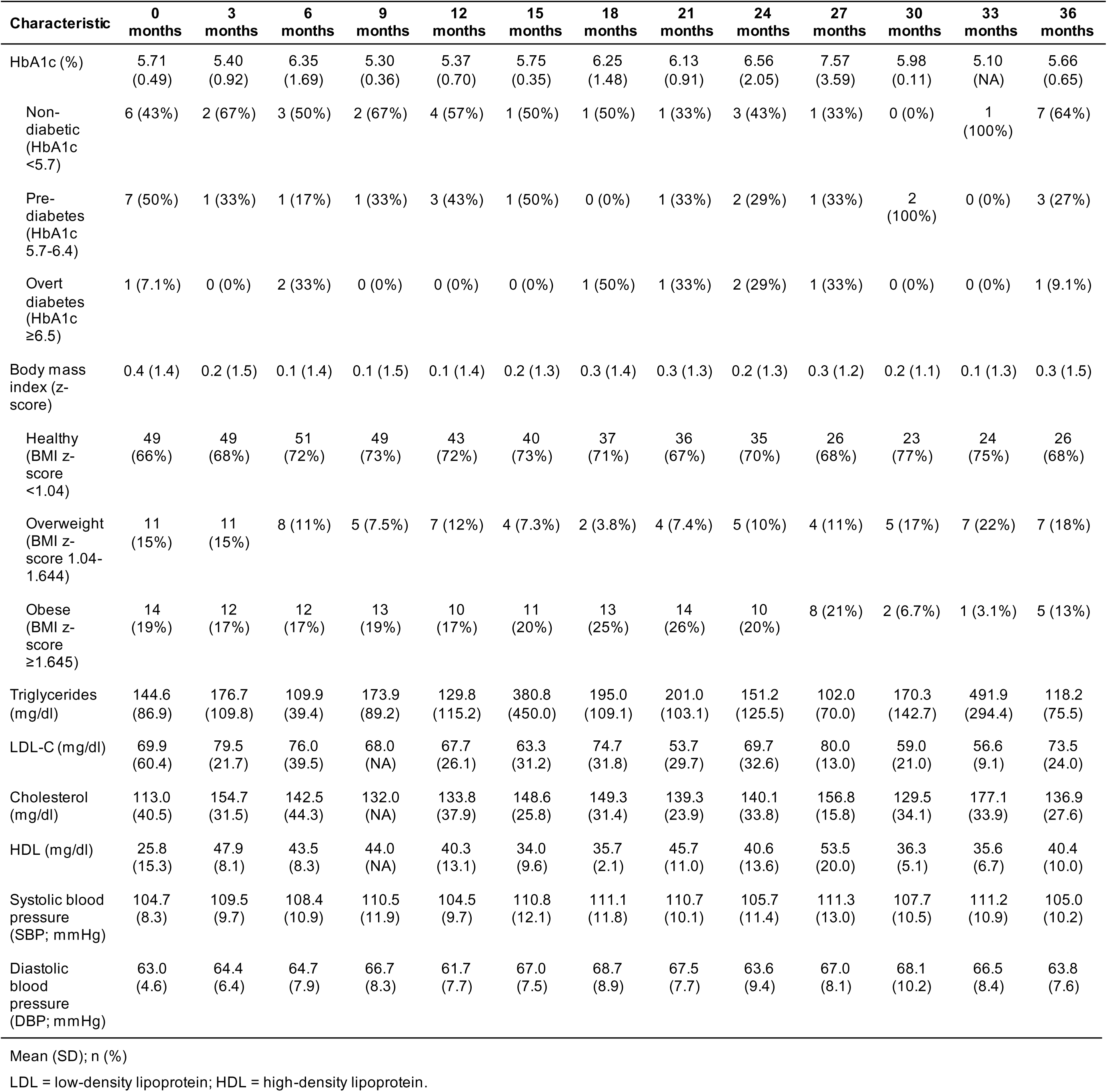
Post-transplant CKM characteristics of pediatric heart transplant recipients.

### Sensitivity analyses

In sensitivity analyses excluding adults who died or were lost to follow-up within 30 days post-HT (N = 810 adults), the IRs of DM2, overweight or obesity, dyslipidemia, and CKD were 28.5, 75.6, 138.1, and 70.4 per 100 person-years, respectively. Similarly, when excluding adults who received simultaneous multiorgan transplants (N = 761 adults), the IRs of DM2, overweight or obesity, and dyslipidemia were 29.3, 74.8, and 129.6 per 100 case-years, respectively.

## DISCUSSION

In a comprehensive analysis of CKM dysfunction at a high-volume transplant center, we found high incidence and prevalence of post-HT DM2, hypertension, dyslipidemia, CKD, and overweight and obesity among both adults and children. Importantly, these comorbidities developed early – within 1-year or less following HT – in a large proportion of individuals, with specific laboratory values over time predictable using GLS models. In a subset of individuals prescribed SGLT2i or GLP1ras post-HT, we observed significant improvements in renal function and BMI, respectively, suggesting an opportunity for early post-HT intervention in modifying the burden of CKM disease trajectory.

Transplant recipients are at increased risk for CKM disease due to a combination of diseases present pre-HT along with exposure to immunosuppressive agents following HT. For the majority of individuals at our center and elsewhere, maintenance immunosuppression consists of calcineurin inhibitors (CNI; e.g., tacrolimus, cyclosporine), anti-proliferatives (e.g., mycophenolate mofetil, azathioprine), and some duration of steroids, with some individuals receiving mTORi (e.g., everolimus, sirolimus) – usually in place of anti-proliferatives – to attenuate development or progression of graft vasculopathy^7,19^. While detailed mechanistic insight into immunosuppression-mediated metabolic dysregulation is outside the purview of this study, each of these therapies has the potential to contribute to CKM dysfunction. CNIs, for example, promote gut uptake of glucose via increased expression of SGLT-1 and result in pancreatic β-cell dysfunction as well as nephrotoxicity that may lead to impaired glycemic control, hypertension, and worsening renal function^20–22^. Steroid-induced hyperglycemia occurs through numerous mechanisms and is a major contributor to the development of post-HT DM2^8,9,19,23^ as well as to weight gain and lipid dysregulation. Finally, mTOR inhibition can result in β-cell failure, impaired insulin sensitivity, reduced glycogen synthesis, and increased HMG-CoA reductase activity, all of which may contribute to DM2 and hypertriglyceridemia^21,24–26^. Collectively, these unique exposures in the transplant population likely contribute significantly to the high incidence and prevalence of CKM dysfunction observed in our study.

In addition to the impacts of immunosuppressive therapies, trends in some markers of cardiometabolic kidney disease may also be explained by other post-HT factors. Predicted post-HT BMI trajectory based on GLS modeling, for example, was characterized by an initial decline in BMI that may reflect early loss of extravascular fluid as cardiac output normalizes or loss of muscle mass during the days and weeks immediately after surgery when some individuals remain relatively inactive and may be acutely malnourished. Thereafter, BMI values steadily increased. In the case of post-HT dyslipidemia, the peak in LDL-C around 3 months post-HT may be explained in part by early statin initiation at most centers. Our findings that HbA1c, triglycerides, and cholesterol are predicted to continue increasing with time after HT may have several potential etiologies. While immunosuppressives may drive early incident or worsening of CKM disease, the development of CKM derangement may sustain or worsen disease further in a vicious cycle of feedback^3,4,27,28^. Furthermore, as cardiac output normalizes following HT, individuals become accustomed to immunosuppressive regimens, and reduction of target immunosuppressive troughs over time – which, in turn, may lead to reduced side-effects – may result in the recrudescence of sedentary lifestyle and poor nutrition that contribute to ongoing CKM dysfunction^29–31^. These data support the notion that early intervention may attenuate this cycle and reduce cumulative CKM disease burden.

The improvements in renal function and BMI observed in a subset of individuals prescribed SGLT2i and GLP1ra have important implications for post-HT management that may mitigate the burden of CKM disease. In recent years, the advent of these drugs has made significant inroads toward addressing CKM disease in the non-transplant population (both adult and pediatric)^32–40^ and several smaller studies in transplant have suggested a role for these drugs after transplant^41–43^. In our study, among 242 adults prescribed SGLT2i and 168 adults prescribed GLP1ras, we observed significant improvements in eGFR and reductions in BMI, respectively. The post-SGLT2i eGFR trajectory in our study exhibited the characteristic curve observed in other studies, with an initial reduction in eGFR until ∼5 months post-initiation, followed by a steady increase in eGFR thereafter^32,33,35^. This initial “dip” in eGFR, followed by eGFR stabilization, is thought to be secondary to the effects of SGLT2i on tubuloglomerular feedback. Although we did not explore the effect of GLP1ras on other CKM endpoints, weight loss has been associated in large-scale studies with significant improvements in cardiovascular disease, kidney dysfunction, and psychiatric illness^44^. While ideal timing of initiation of these therapies post-HT remains unclear, the significant burden of CKM dysfunction observed within 1-year post-HT in our study supports early intervention.

While the overall burden of CKM disease in pediatric HT recipients was lower in our study, it remains higher than the non-HT population^45–47^. Similar to adults, it is likely that immunosuppressive regimens following solid-organ transplantation contribute to this phenomenon. Additionally, the same non-transplant (“traditional”) risk factors contributing to the childhood obesity epidemic may be influential as well^45,46^. These risks may be compounded by data suggesting that nearly two-thirds of pediatric HT recipients may not be on statin therapy^48,49^. The dyslipidemia burden may be particularly relevant in light of increasing understanding that cumulative exposure to LDL-C and other lipids may impact future cardiovascular risk^50^. Importantly, despite the overall lower incidence and prevalence of CKM burden in children as compared to adults in our study, prior work has shown that cardiometabolic disease increases the risk of graft vasculopathy and graft failure in pediatric HT recipients^47^. As SGLT2i, GLP1ras, and other novel therapeutics are increasingly being studied in the pediatric population, early intervention in children who undergo HT may attenuate long-term consequences of CKM dysfunction.

It is important to frame our findings in the context of several limitations. First, not all 944 adults and children studied had complete CKM data due to (1) a small portion of individuals who were transplanted elsewhere and transferred their post-HT care to Vanderbilt later, and (2) laboratory protocols that evolved over time such that not all relevant CKM values were available at all time points of interest for all individuals. Our use of GLS models with AR(1) helps to mitigate these issues by extrapolating missing data using data from surrounding time points in order to construct laboratory trajectories over time. Second, because incidence rates were calculated based on the time post-HT at which individuals first met the outcome of interest (e.g. eGFR <45 mL/min/1.73 m^2^), we did not account for subsequent improvements in laboratory values that may have occurred thereafter. Third, as with all EHR-based studies, it is possible that miscoding of ICD diagnoses codes may have impacted the accuracy of our results. To minimize this possibility, we prioritized study of captured laboratory values related to diagnoses (e.g., HbA1c, lipid profiles, eGFR, etc.) rather than relying on ICD9 or ICD10 codes alone. Finally, our study is limited to a single-center experience, which may not be generalizable to all transplant centers.

In a large population of adult and pediatric heart transplant recipients at a high-volume center, we observed significant CKM disease that developed early post-HT. Novel drug therapies may substantially mitigate CKM burden and improve post-transplant outcomes. Further work is needed to guide timing of initiation of these therapies and to explore their impacts on longer-term outcomes including graft vasculopathy and mortality.

## Data Availability

All data produced in the present work are contained in the manuscript

## ACKNOWLEDGMENTS

Dr. Amancherla acknowledges Dr. Ravi V. Shah for his support and mentorship.

